# Distinct neural alterations in schizophrenia and autism: A meta-analysis of social cognition and emotion processing

**DOI:** 10.1101/2024.10.11.24315338

**Authors:** Mélanie Boisvert, Florence Pilon, Laurent Mottron, Stéphane Potvin

**Author notes:** **Co-corresponding authors**: Stéphane Potvin, PhD; Centre de recherche de l’Institut Universitaire en Santé Mentale de Montréal; 7331 Hochelaga; Montreal, Que, Canada; H1N 3V2; / Laurent Mottron, MD, PhD, Hopital Rivière des Prairies, 7070 Bvd Perras, Montréal, Que, Canada, H1E1A4. LM and SP made equal contributions to the manuscript.

## Abstract

**Objective:** Two conditions alter socio-communicative behaviors in humans: autism and schizophrenia. However, it is not well-known if these disorders share the same neural alterations during socio-emotional tasks. The main objective was to examine neural alterations in autism and schizophrenia during emotional and social cognition tasks. Our second objective was to determine if these alterations were common or distinct between disorders.

**Methods:** Functional neuroimaging studies using an emotional or a social cognition paradigm in schizophrenia or autism were queried from three databases. We selected articles if they reported whole brain coordinates of different activations between autism/schizophrenia participants and non-clinical controls. Using SDM, we analyzed the coordinates of brain activity differences between case and control groups, categorized by diagnosis and paradigm.

**Results:** The meta-analysis aggregated 104 studies in schizophrenia and 80 studies in autism spectrum disorder. During emotional tasks, individuals with autism showed reduced activity in the left amygdala, while those with schizophrenia showed reduced activity in the right inferior frontal gyrus and the median cingulate gyrus. During social cognition tasks, alterations in both conditions did not survive corrected statistical thresholds. No spatial conjunction was observed between the alterations seen in each disorder during both emotional/social cognition tasks at both corrected and uncorrected thresholds.

**Conclusions:** These results suggest that the emotional processing in autism and schizophrenia in adulthood are characterized by alterations of bottom-up and top-down mechanisms of the emotional network, respectively. It should encourage the pursuit of functional neuroimaging studies on emotion processing using machine learning to differentiate these two conditions.

## 1. Introduction

Two major conditions alter socio-communicative behaviors in humans: autism and schizophrenia. The term “autism” was borrowed from schizophrenia, and the two conditions were initially confused, as evidenced by the name of the journal “autism and childhood schizophrenia” until 1979 (1). The evolution of classifications and knowledge has not clarified the phenotypic, neurocognitive, mechanistic, genetic, and epidemiological relationship between the two conditions. From a clinical point of view, we assist in the last decade to a gradual abandoning of features traditionally supporting the distinction of autism spectrum disorder (ASD) from schizophrenia, with a gradual equalization of sex ratio (2) and an acceptance of late diagnosis (3) in ASD. Behavioral ‘’traits’’ instruments measuring social competence and associated emotions struggle to differentiate them (4). Both being identified as ‘’spectrums’’ without inner subtyping since DSM-5, their boundaries with normalcy are imprecise: autistic traits (5), and schizoid features (6) are found in non-clinical population, favoring a semiological overlap between the least prototypical presentations of each condition. Their accepted heterogeneity, specifically for ASD but a pervasive issue also in schizophrenia research contributes to render their distinction fuzzy. In the case of ASD, diagnostic criteria offer an unlimited overture to co-morbidity (DSM-5) supporting claims between 1% to 61% of comorbidity between the two conditions (7), which is vulnerable to trivial interpretations based on the practical impossibility to firmly separate their presentation, particularly in adults. At the neurocognitive level, socio-emotional impairment is shared between the two conditions, particularly for theory of mind, alexithymia and reduced emotional expression (8,9). Genetic studies of common variants have identified numerous similarities between these two conditions (10), supporting claims that advocate for their potential integration into a continuum, both towards each other and towards normalcy (11,12). Multiple copy number variants (CNVs) produce monogenic pictures, or intermediate pictures between the two conditions, or result in a pleiotropic disease where the two conditions are separately represented, as in the case of CNVs at 22q11 (13). At the epidemiologic level, the blurring of their distinction is favoured unequally by the temporal evolution of each of the two conditions, with prevalence of autism constantly increasing, which is not the case for schizophrenia (14).

Specifying biomarkers to separate ASD and schizophrenia has been yet disappointing due to their modest replicability in ASD (15) as well as in schizophrenia (16). Understanding how distinct are these conditions has, however, considerable consequences on clinical identification, therapeutic support as well as on research strategy (17). Some have argued that the success of the ‘’transdiagnostic revolution’’ in psychiatry (18) may be limited to certain dimensions as anxiety and depression (19) and may spare the relatively autonomy between ASD and schizophrenia, supporting the psychosis-autism diametrical model (20). Recent demonstration of developmental and phenotypic differences in the socio-emotional component of the two conditions (21,22) leads to reconsider the actual level of overlap of the neural correlates of social cognition and emotion processing in autism spectrum and schizophrenia, the major areas of symptoms overlap between the two disorders (23–25). In terms of global performance, autism and schizophrenia seem to share socio-emotional and socio-cognitive deficits that appear very similar. However, some authors have argued that the processes underlying these deficits appear to differ markedly between autism and schizophrenia, suggesting the existence of potential neurobiological differences between the two disorders (26,27). More than a decade ago, Sugranyes et al. (28) performed a meta-analysis of 33 functional neuroimaging studies on social cognition in ASD and schizophrenia, which revealed that both disorders shared common alterations in the medial prefrontal cortex and the superior temporal sulcus, while between-group differences were observed in the amygdala, somatosensory cortex and thalamus (28). In the last decade, a very large number of schizophrenia and autism studies have been published using social cognition or emotional paradigms, raising the need to update the previous meta-analysis.

## 2. Materials and methods

### 2.1 Literature search

Records were queried from three different databases on August 2023: Web of Sciences, Pubmed and EMBASE, which host all the records from Web of Science Core Collection, MEDLINE, KCI-Korean Journal Database, Current Contents Connect, SciELO Citation Index and Preprint Citation Index. The search query included: [(“neuroimaging” OR “fMRI” OR “functional magnetic resonance imaging”) AND (“emotion” OR “social-cognition” OR “theory of mind” OR “mentalize” OR “empathy” OR “pain” OR “facial” OR “belief” OR “judgement” OR “irony”)]. For the autism search, the terms [(“autism OR “asperger”)] were added whereas for the schizophrenia research [(Schizophrenia)] was specified.

### 2.2 Selection criteria

Studies were first selected if they met the following criteria: (i) included a measurement of brain activity (positron emission tomography (PET), single-photon emission computed tomography or functional magnetic resonance imaging [fMRI]) during an emotional or social cognition task, (ii) included an ASD or schizophrenia group; (iii) included a non-clinical control group. Excluded were studies that: (i) had clinical groups younger than 18 years or older than 40 years (to match more closely the two clinical groups since younger cohorts are studied in ASD and older cohorts in schizophrenia), (ii) analyzed brain activity during rest, (iii) reported results for predefined regions of interest, (iv) did not perform between-group comparison. Studies were reviewed by two researchers (MB, FP), inclusion/exclusion criteria were evaluated by consensus. We followed the “*Preferred Reporting Items for Systematic Reviews and Meta-Analyses*” (PRISMA) guidelines (29).

### 2.3 Data extraction

Data extraction was completed by two researchers (MB, FP). For each study included, the following information was extracted: (i) first author and year of publication; (ii) number of participants and demographics (age, sex ratio); (iii) ASD or schizophrenia phenotype; (iv) positive and negative symptoms of psychosis; (v) chlorpromazine dose equivalents; (vi) smoothing level; (vii) scanner type (MRI field strength, fMRI vs PET); (viii) voxel size; (ix) time of repetition; (x) type of task. The classification of tasks into social cognition or emotion domains was based on previous meta-analyses performed in ASD and/or schizophrenia (24,28), and was done by two researchers (MB and FP). If there was an affective component in the task, the study was classified as emotional (passive view of angry faces would be emotional, whereas a false-belief task would be classified as social cognition). For the clinical phenotype, we extracted the percentage of participants with a diagnosis of autism (vs Asperger, etc.) and the percentage of participants with a diagnosis of schizophrenia (vs schizoaffective or other psychotic disorder). Inter-judge agreement for task classification and ASD phenotype classification (% of autism diagnosis) was 94%, and 97% for schizophrenia phenotype classification. Discrepancies were resolved by consensus. Antipsychotic dose was calculated using chlorpromazine equivalents (30,31). Positive and negative symptom scores from the Scale for Assessment of Positive and Negative Symptoms were converted to Positive and Negative Syndrome Scale scores using equations implemented at http://converteasy.org.

### 2.4 Coordinate-based meta-analysis

A voxel-wise meta-analysis was performed using the SDM-PSI version 6.11 (32). The SDM-PSI is a software using peak coordinates and their *t*-values as reported from the original studies, to impute, for each study, multiple effect-size maps (Hedges’ g) of contrast results (increased and decreased activations). Maps are then combined in a standard random-effects model considering sample size, intra-study variability and between-study heterogeneity (33). Multiple imputations are pooled using Rubin’s rules (32). The familywise error rate of the results is calculated using a subject-based permutation test (34). SDM-PSI uses MetaNSUE (35) to estimate the maximum likely effect size within the lower and upper bounds of possible effects sizes for each study separately and adds realistic noise (32,34).

### 2.5 Meta-analytic procedure

Main meta-analyses were performed to assess differences in brain activity between psychiatric patients and non-clinical controls during emotional and social cognition paradigms. Analyses were performed in the sets of studies on schizophrenia and ASD separately. Residual heterogeneity of included studies was examined using the *I*^2^ statistic (*I*^2^ >50 % indicates significant heterogeneity). Potential publication bias was assessed via a meta-regression analysis of the effect size by its standard error (36). To assess the robustness of the results, we performed sensitivity analyses by sequentially removing each study and rerunning the analysis using the regional effect size estimates. Results are reported corrected for multiple comparisons using threshold-free cluster enhancement (TFCE; 1 000 permutations), with a corrected *p*<0.05 and cluster extent of 20 voxels. Analyses were also performed using uncorrected thresholds (peak: *p*<0.001 and cluster threshold >50 voxels), as in several fMRI meta-analyses (35,37,38). Since social cognition tasks were heterogeneous, we decided to perform secondary meta-analyses in both disorders for studies specifically using mental attribution paradigms (false-belief, theory of mind). These kinds of paradigms were used in a sufficient number of social cognition studies to enable use to perform secondary meta-analyses.

### 2.6 Conjunction and difference analyses

To investigate the co-occurrence of functional abnormalities in schizophrenia and ASD, we used the multimodal analysis in SDM-PSI. This conjunction analysis involved overlaying thresholded meta-analytic results maps of schizophrenia brain activity with maps of ASD alterations. This comparison aimed to identify areas of spatial convergence in results across both disorders. The conjunction analysis is the standard approach in SDM-PSI to identify common alterations in multiple disorders, and has been used in several fMRI meta-analyses (39,40).

Potential differences between disorders were examined using linear regression analyses, as in several MRI meta-analyses using SDM-PSI (39,41). These analyses were only performed on the paradigm(s) revealing significant results surviving TFCE correction *within* disorders.

### 2.7 Meta-regressions and sub-analyses

Meta-regression and sub-analyses were performed to assess the moderation effect of demographic and clinical variables including age, sex ratio, percentage of autism phenotype, percentage of schizophrenia diagnosis, chlorpromazine equivalents, positive and negative symptoms of schizophrenia. The moderation effects of neuroimaging parameters (smoothing level, voxel size, time repetition and scanner type such as fMRI field strength and fMRI vs PET) were investigated. Meta-regression analyses were done using *Comprehensive Meta-analysis* 2 (42), and were restricted to brain regions that were found to be altered using TFCE correction in the meta-analyses. In these analyses, the dependent variables were the effect sizes for each study for each cluster and the predictors were the confounding variables.

### 2.8 Replicability of results

As small differences may be observed between SDM-PSI and other meta-analytical approaches. GingerALE is based on the spatial convergence of peaks across studies and only considers significant between-group results. On the other hand, SDM-PSI is based on effect sized estimates and considers both positive and negative results in the analyses. Therefore, we also ran the above-described analyses using the ALE method (see Supplementary material).

## 3. Results

### 3.1 Included studies

The initial searches yielded 3 788 articles in schizophrenia and 2 836 studies in ASD (Supplementary Figure 1 and 2 and Supplementary Table 1). One hundred and four studies were included comprising 121 contrasts in 2,381 participants with schizophrenia (mean age=32.7±4.3; 34% female; only 4% with schizo-affective disorder or other psychotic disorders) compared to 2,488 non-clinical controls. Eighty studies were also included comprising 86 contrasts representing 1 561 participants with ASD (mean age=26.9±4.8; 17% female; 58% of autism) compared to 1 618 neurotypical controls. Emotional tasks used social stimuli in 72% of schizophrenia studies and 88% of ASD studies (see Supplementary Tables 2-6). Noteworthy, the ratio of paradigm types did not differ between disorders. In the case of social cognition, there were no differences in the ratio of inference tasks (theory of mind task, false belief task, reading the mind in the eyes, etc.) (X^2^(1, 65)=0.103, p=0.749) versus other social cognition tasks (eye gaze processing, approachability judgement, humor processing, etc.) (X^2^(1, 192)=0.005, p=0.945). As for emotional tasks, there were no differences between disorders in the ratio of explicit/implicit emotional tasks (X^2^(1, 155)=0.001, p=0.973) (Supplementary Table S6).

### 3.2 Meta-analysis of emotional studies in schizophrenia

At a TFCE corrected level (p<0.05), schizophrenia patients displayed reduced brain activity compared to controls in the right (triangular) inferior frontal gyrus (IFG) and the right median cingulate gyri during emotional paradigms (Table 1, Figures 1A, 1B and 2). At an uncorrected threshold (p<0.001), no other brain regions were found to be altered in schizophrenia (see Supplementary Table 7).

**Figure 1.**
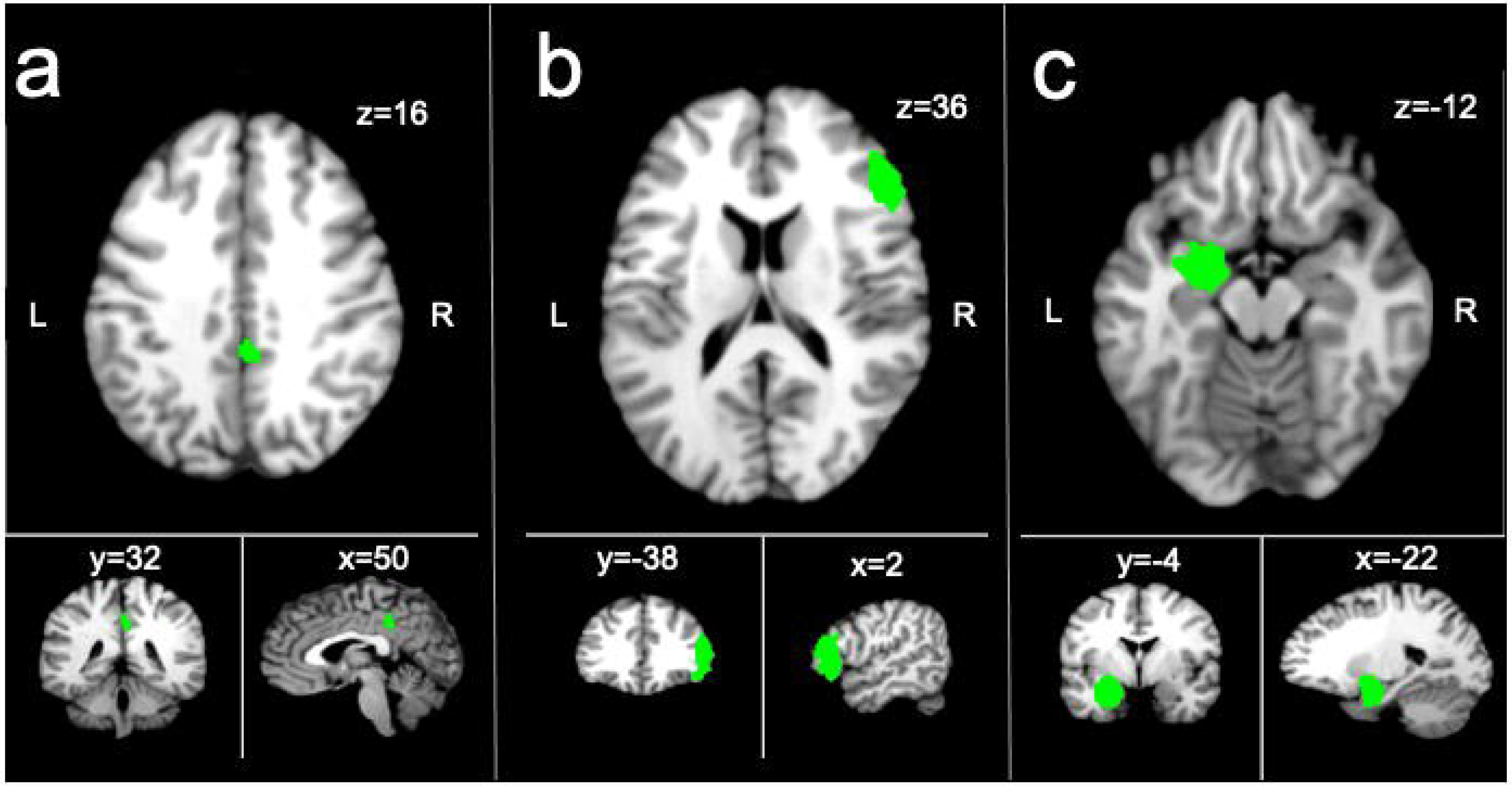
Brain regions displaying lower activations in clinical groups compared to non-clinical controls at a TFCE corrected threshold (p<0.05) in an SDM-PSI meta-analysis on emotional studies. A=right median cingulate in schizophrenia; B=right inferior frontal gyrus in schizophrenia; C=amygdala in autism spectrum disorder.

**Table 1.**
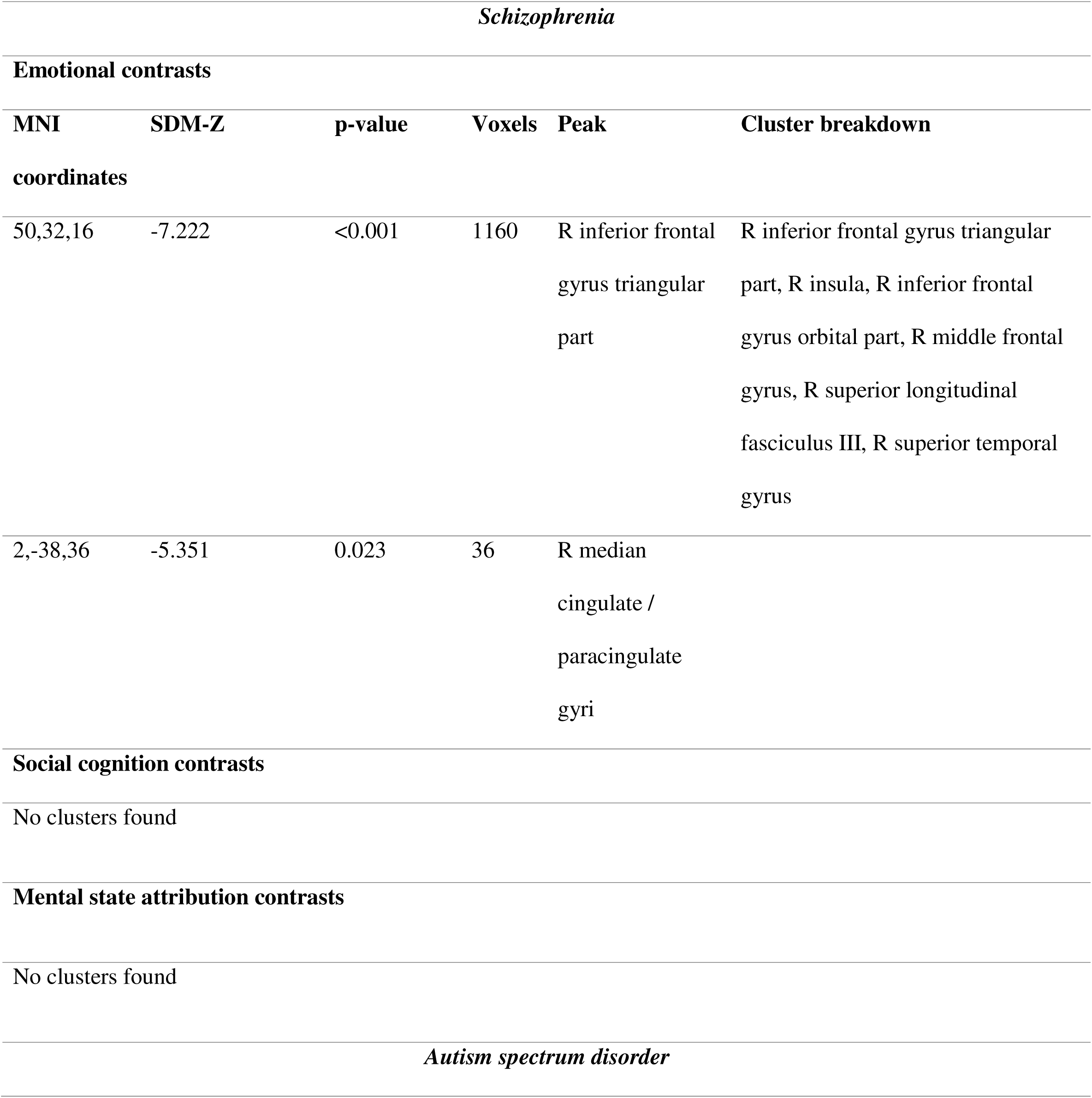
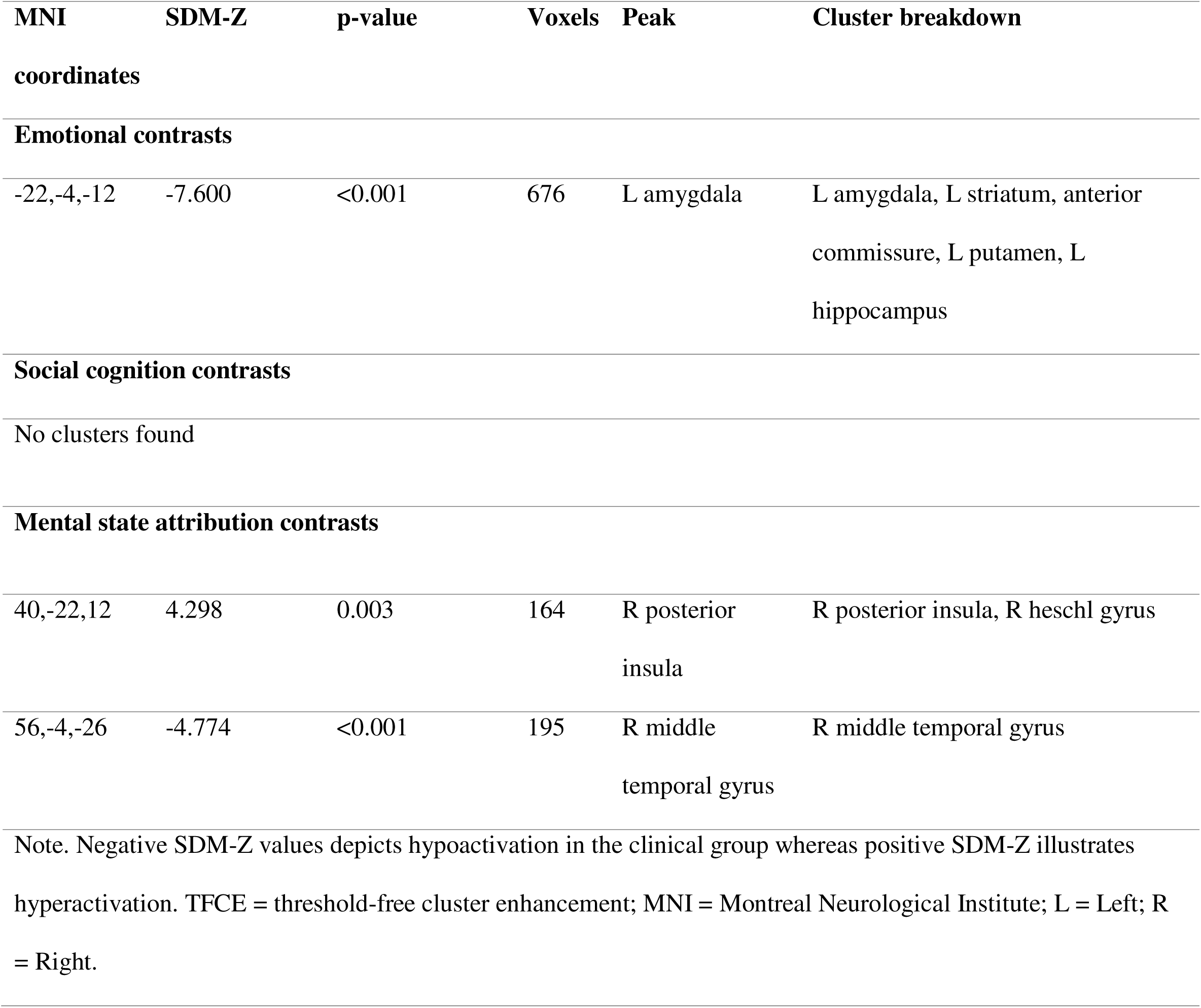
TFCE corrected results from SDM-PSI displaying altered activation in clinical groups compared to non-clinical controls.

There was very low heterogeneity (*I*^2^=0.32-2.4%) and no evidence of publication bias (both ps>0.4). The results were robust as no study affected the meta-analytical estimates by more than 5 %.

We found no association between chlorpromazine dose, age, sex, psychotic symptoms, percentage of schizophrenia diagnosis or scanner parameters (all p-values>0.1) and the alterations found in the right IFG and the right median cingulate gyri (Table 2 and 3).

**Table 2.** Meta-regressions.

| Moderators | Slope | Confidence interval<br>(95%) | p-value |
| --- | --- | --- | --- |
| Schizophrenia |  |  |  |
| *R Inferior Frontal Gyrus* |  |  |  |
| Age | 0.0029 | -0.0188 to 0.0245 | 0.7957 |
| Sex ratio (% of females) | -0.0021 | -0.0076 to 0.0035 | 0.4666 |
| Chlorpromazine dose | 0.0002 | -0.0003 to 0.0008 | 0.4426 |
| Positive symptoms | -0.0022 | -0.0276 to 0.0233 | 0.8673 |
| Negative symptoms | 0.0003 | -0.0223 to 0.0228 | 0.9813 |
| Diagnosis (% of schizophrenia patients) | 0.0001 | -0.0053 to 0.0054 | 0.9827 |
| Smoothing level | 0.0178 | -0.0493 to 0.0850 | 0.6027 |
| Voxel size | 0.0006 | -0.0041 to 0.0053 | 0.8036 |
| Time repetition | 0.0000 | -0.0002 to 0.0002 | 0.9748 |
| *R Median Cingulate Cortex* |  |  |  |
| Age | -0.0030 | -0.0351 to 0.0291 | 0.8560 |
| Sex ratio (% of females) | 0.0043 | -0.0041 to 0.0127 | 0.3141 |
| Chlorpromazine dose | 0.0001 | -0.0006 to 0.0008 | 0.7346 |
| Positive symptoms | -0.014 | -0.0346 to 0.0318 | 0.9354 |
| Negative symptoms | -0.0022 | -0.0336 to 0.0291 | 0.8883 |
| Diagnosis (% of schizophrenia patients) | -0.0012 | -0.0162 to 0.0138 | 0.8714 |
| Smoothing level | 0.0246 | -0.0606 to 0.1099 | 0.5715 |
| Voxel size | 0.0002 | -0.0047 to 0.0051 | 0.9429 |
| Time repetition | -0.0000 | -0.0002 to 0.0002 | 0.8178 |
| Autism spectrum disorder |  |  |  |
| *L Amygdala* |  |  |  |
| Age | 0.0091 | -0.0116 to 0.0298 | 0.3892 |
| Sex ratio (% of females) | 0.0006 | -0.0039 to 0.0050 | 0.8075 |
| Diagnosis (% of autism) | 0.0006 | -0.0017 to 0.0029 | 0.5904 |
| Smoothing level | -0.0074 | -0.0820 to 0.0671 | 0.8551 |
| Voxel size | 0.0007 | -0.0045 to 0.0059 | 0.7923 |
| Time repetition | 0.0000 | -0.0001 to 0.0001 | 0.9501 |
| *R posterior insula* |  |  |  |
| Age | -0.006 | -0.049 to 0.037 | 0.794 |
| Sex ratio (% of females) | 0.001 | -0.008 to 0.011 | 0.784 |
| Diagnosis (% of autism) | 0.0006 | -0.005 to 0.006 | 0.814 |
| Smoothing level | -0.034 | -0.192 to 0.124 | 0.674 |
| Voxel size | 0.003 | -0.006 to 0.013 | 0.502 |
| Time repetition | 0.0001 | -0.0002 to 0.0005 | 0.529 |
| *R middle temporal gyrus* |  |  |  |
| Age | -0.011 | -0.055 to 0.032 | 0.61 |
| Sex ratio (% of females) | 0.0001 | -0.009 to 0.009 | 0.981 |
| Diagnosis (% of autism) | -0.0005 | -0.006 to 0.005 | 0.863 |
| Smoothing level | -0.018 | -0.174 to 0.138 | 0.862 |
| Voxel size | 0.002 | -0.008 to 0.011 | 0.736 |
| Time repetition | 0.00004 | -0.0003 to 0.0004 | 0.822 |
| Note. R = Right; L = Left |  |  |  |

**Table 3.**
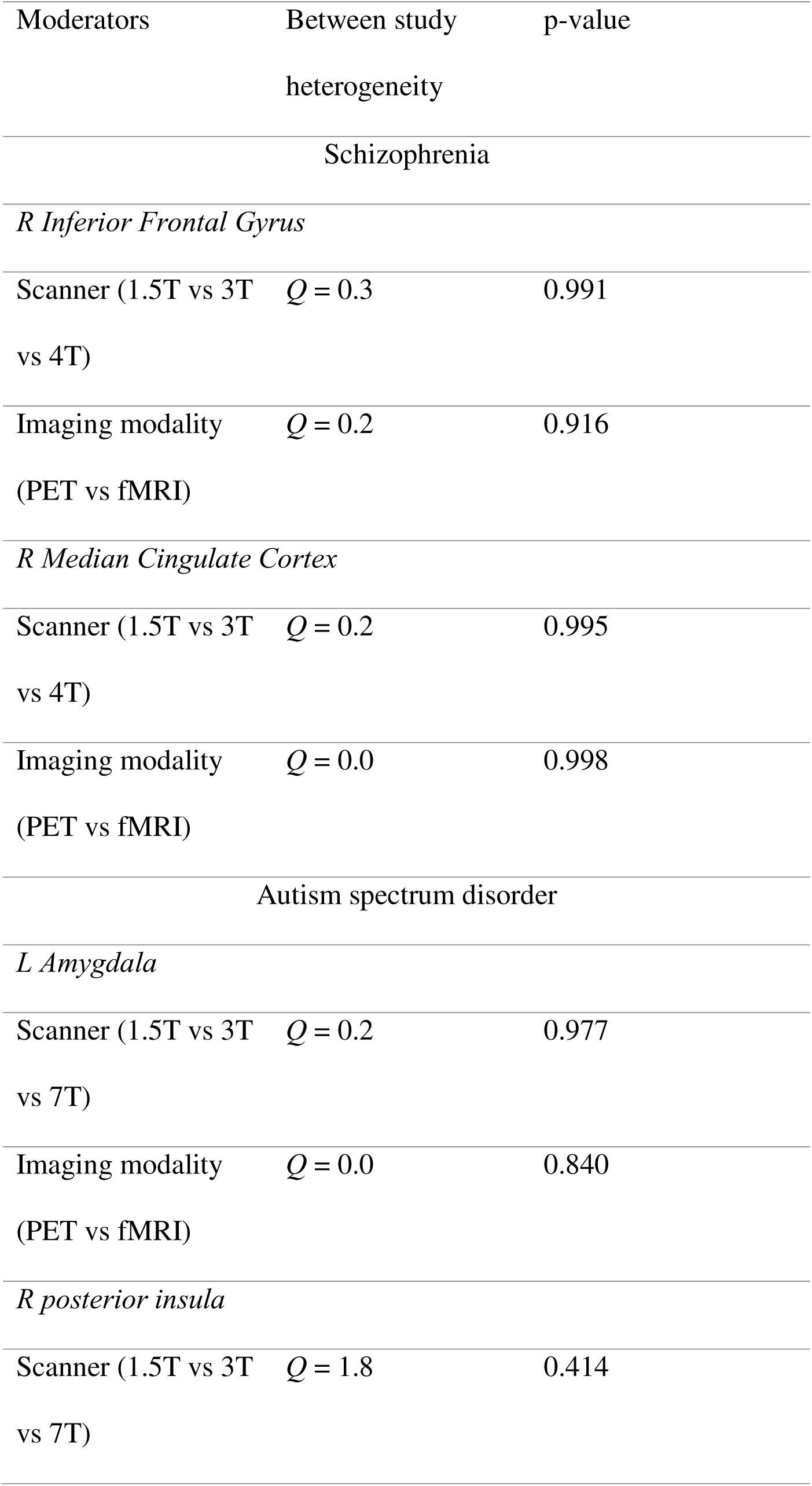
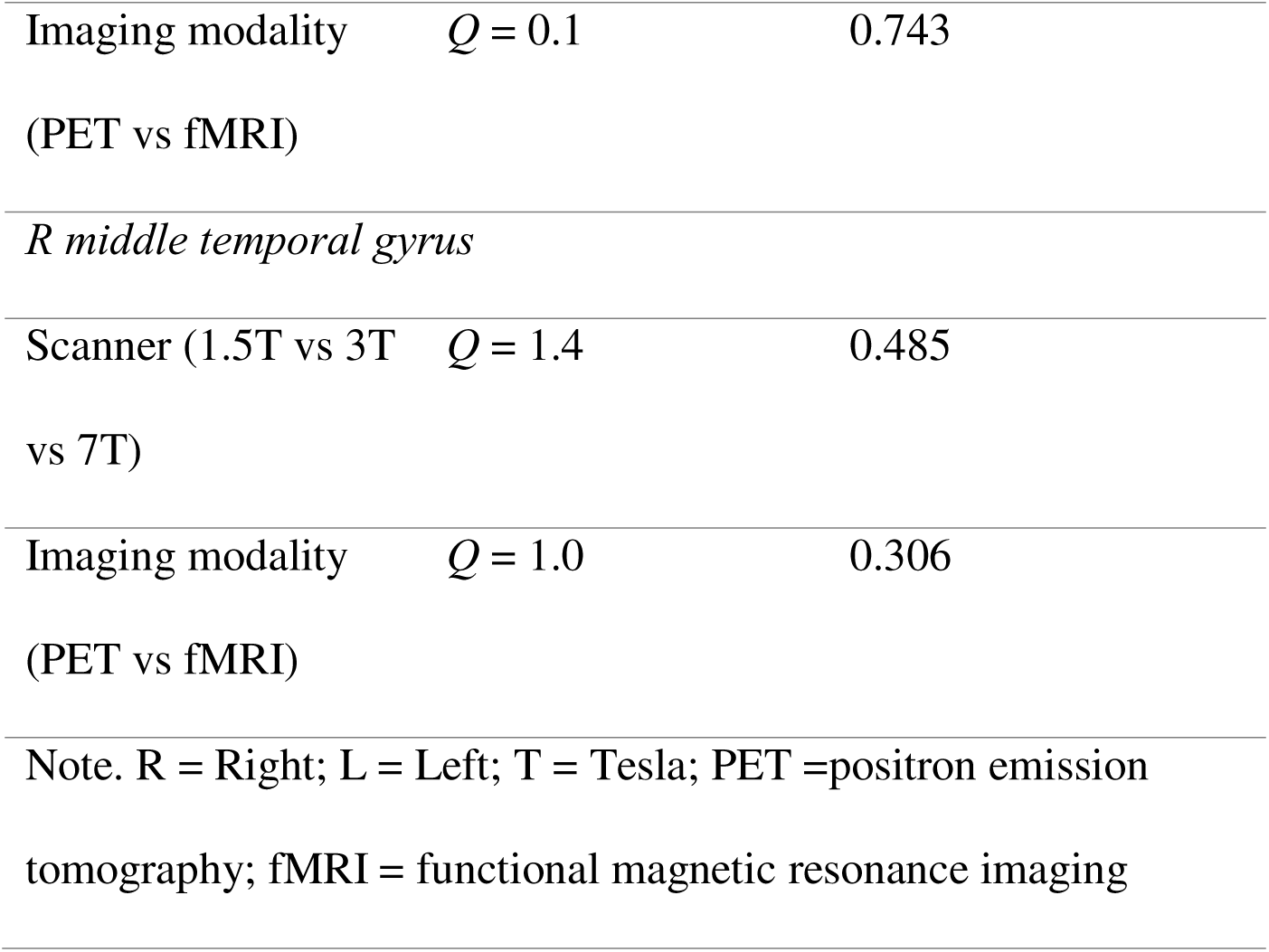
Subanalyses.

### 3.3 Meta-analysis of social cognition studies in schizophrenia

Using a stringent statistical threshold (TFCE p<0.05), no differences were found in brain activity during social cognition paradigms between schizophrenia and non-clinical controls. At an uncorrected threshold (p<0.001), reduced activity was observed in the right middle temporal gyrus in schizophrenia (see Supplementary Table 7).

### 3.4 Secondary meta-analysis on mental state attribution studies in schizophrenia

No clusters were found both at a corrected threshold (TFCE p<0.05) and an uncorrected threshold (p<0.001).

### 3.5 Meta-analysis of emotional studies in ASD

At a corrected threshold (TFCE p<0.05), one cluster displayed lower activation in the group with a diagnosis of ASD compared to neurotypical controls, which was the left amygdala (Table 1, Figures 1C and 2). The ASD group also displayed reduced activations in the left inferior frontal gyrus (opercular part), the left fusiform gyrus and the right para-hippocampal gyrus when using an uncorrected threshold (see Supplementary Table 8).

**Figure 2.**
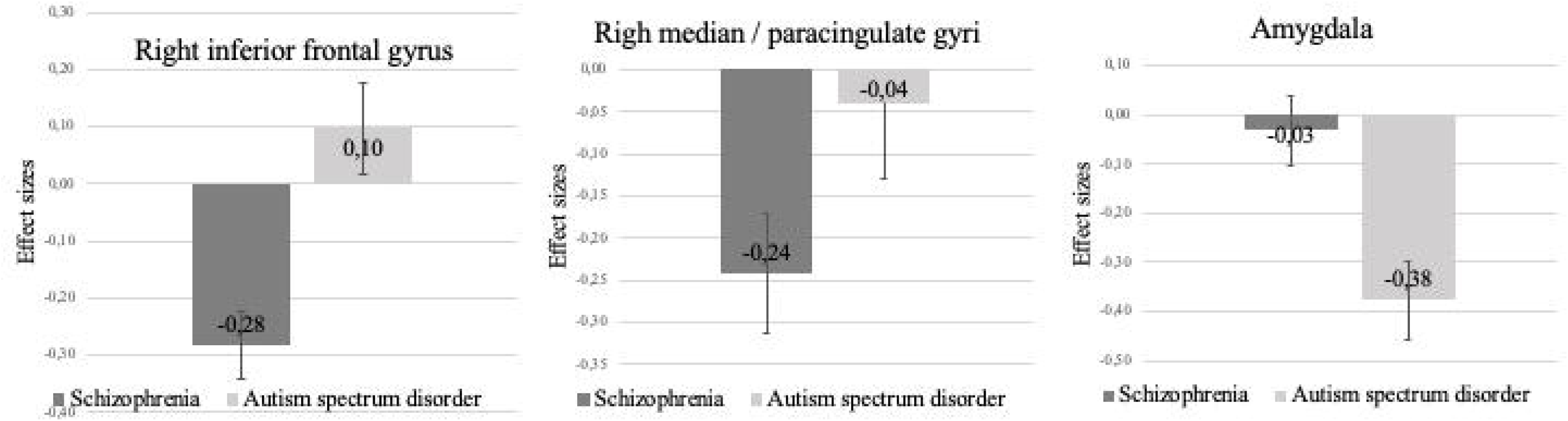
Between-group comparisons performed using linear regressions. Note: The error bars are standard deviations.

There was no evidence of heterogeneity (*I*^2^=0.20%) or publication bias (p=0.559). The amygdala results were robust as no study affected the meta-analytical estimates by more than 5 %.

We found no association between phenotype, age, sex ratio, smoothing level, voxel size, type of scanner, or time repetition (p-values>0.1) and neural alterations found in the amygdala (Table 2 and 3).

### 3.6 Meta-analysis of social cognition studies in ASD

Using a stringent statistical threshold (TFCE p<0.05), no differences were found in brain activity during social cognition paradigms between ASD and controls. At an uncorrected threshold, reduced activations were found in the ASD group in the right superior temporal gyrus (see Supplementary Table 8).

### 3.7 Secondary meta-analysis on mental state attribution studies in ASD

In the secondary meta-analysis on mental state attribution studies, using a stringent threshold (TFCE p<0.05), two clusters were found to be significantly altered in ASD compared to non-clinical controls. Increased activation was found in a cluster located in the right posterior insula (Table 1, Figure 3A), while decrease activation was found in the right middle temporal gyrus (Table 1, Figure 3B). At an uncorrected threshold (p<0.001) an additional cluster of decreased activation was found in the left superior frontal gyrus, medial part.

**Figure 3.**
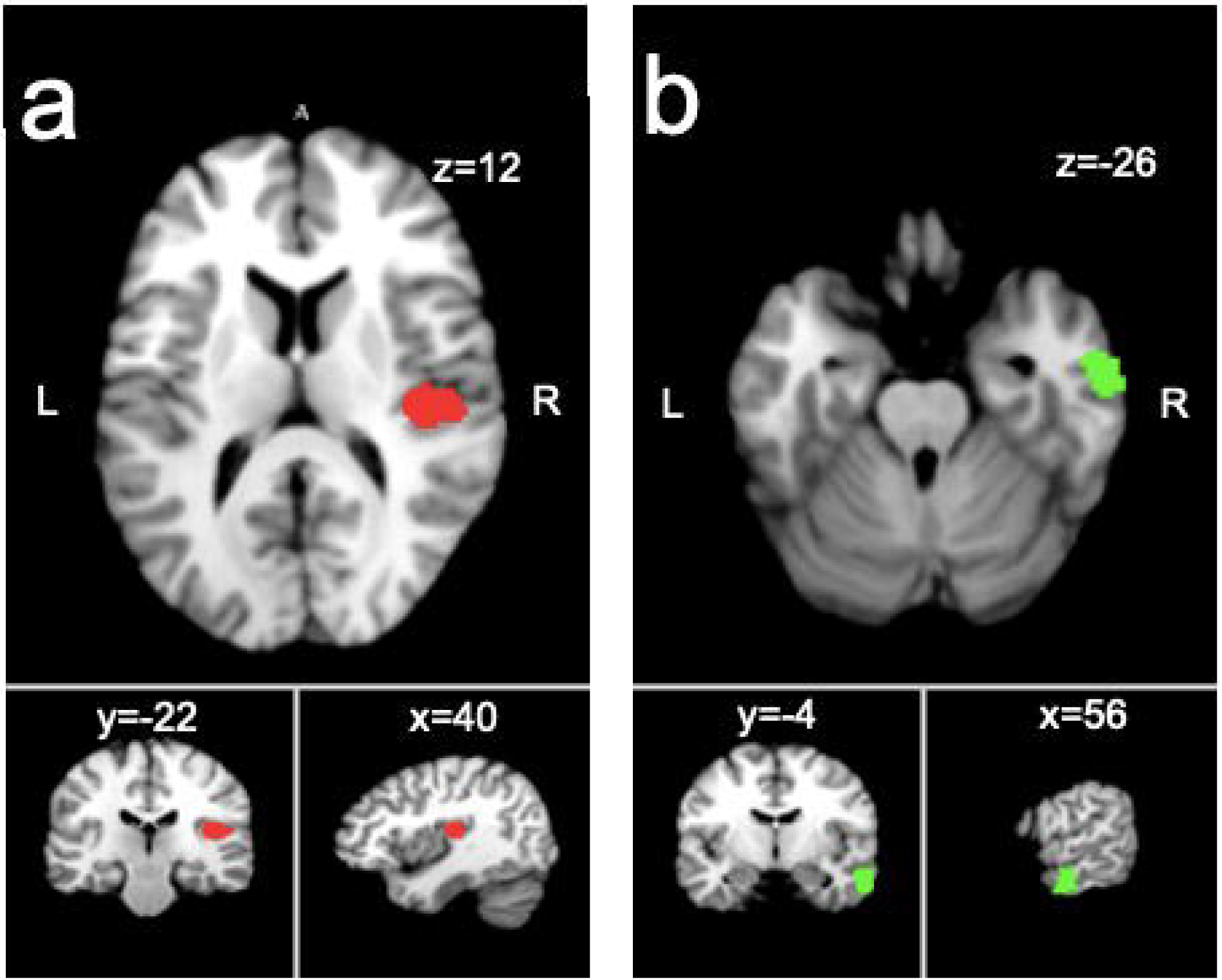
Brain regions displaying altered activations in autism spectrum disorder compared to non-clinical controls at a TFCE corrected threshold in an SDM-PSI meta-analysis on mental state attribution studies. A = increased activation of the posterior insula; B = decreased activation of the middle temporal gyrus.

There was low heterogeneity (*I*^2^=1.19-4.79%) and no evidence of publication bias (all ps>0.5). The results were robust as no study affected the meta-analytical estimates by more than 5 %.

We found no association between phenotype, age, sex ratio, smoothing level, voxel size, type of scanner, or time repetition (p-values>0.1) and neural alterations found in the right posterior insula or right middle temporal gyrus (Table 2 and 3).

### 3.8 Conjunction and difference analyses

For, emotional, social cognition and mental state attribution paradigms, no overlapping clusters were found at a stringent corrected threshold (TFCE p<0.05) and at an uncorrected threshold (p<0.001) between schizophrenia and ASD.

The linear regression analyses revealed significant differences *between* schizophrenia and ASD in the right IFG (Z=6.8; p=0.0001), the median cingulate (Z=3.6; p=0.0003) and the amygdala (Z= -6.1; p=0.0001) during emotional paradigms (Supplementary Figure 3). In our secondary meta-analysis on mental state attribution, differences *between* schizophrenia and ASD were also noted in the right posterior insula (Z=4.7; p<0.001) and the right middle temporal gyrus (Z=5.3; p<0.001).

### 3.9 Replicability of results

Our main findings (right IFG alteration in schizophrenia and left amygdala alteration in ASD during emotional paradigms; and lack of conjunction between disorders) were replicated using ALE (see Supplementary Tables 9 and 10).

## 4. Discussion

Considering that schizophrenia and ASD both comprise socio-cognitive and socio-emotional phenotypical manifestations, we performed a meta-analysis of 184 functional neuroimaging studies aggregating activation differences between clinical and non-clinical individuals during emotional and social cognition tasks. We found reduced activity in the left amygdala in ASD, and reduced activity in the right IFG and the right median cingulate gyrus in schizophrenia during emotional tasks. Alterations in temporal regions were observed during social cognition tasks, but only when using uncorrected statistical thresholds. In mental state attribution studies specifically, additional alterations were found in the ASD group in the right posterior insula and the right middle temporal gyrus. The conjunction analyses revealed no neural alterations common to both disorders.

Socio-emotional processing involves complex neural networks encompassing brain regions involved in face perception (fusiform gyrus), affective responding (amygdala, hippocampus), self-other differentiation (medial prefrontal cortex, precuneus, and temporo-parietal junction), and cognitive control (ventro-lateral prefrontal cortex) (43). This hierarchically organized network involves bottom-up mechanisms relaying stimuli information from perceptual regions to limbic regions and to self-processing regions, and top down-down mechanisms of emotion regulation mediated by executive regions (43).

In ASD, a robust reduction of the left amygdala activity during emotion processing was observed at a stringent correction level. This result is coherent with the amygdala hypothesis of autism formulated by Baron-Cohen et al. (44) and recently updated (45). This theory is based on the observations that the amygdala is involved in threat detection (fear) and in face perception and attention (eye gaze) (45). Damage to the amygdala in animals significantly impairs prosocial behavior; however, the amygdala subregions involved in lesion-induced behaviors do not perfectly coincide with the regions of activity differences indicated by case-control human neuroimaging studies (45). Our results indicate that the bottom-up mechanisms involved in the early detection of emotional stimuli underlie the socio-emotional positive and negative manifestations associated with ASD. Using an uncorrected statistical threshold, other brain regions (fusiform and para-hippocampal gyrus), which are known to be involved in socio-emotional processing (46), were found to be altered in ASD.

Schizophrenia is characterized by emotional blunting and emotion regulation difficulties, and deficits in emotional recognition (47). In schizophrenia, the activity in the right IFG and the median cingulate gyrus was reduced during the emotional tasks. Robust evidence from a meta-analysis of 225 studies has shown that the right IFG plays a crucial role in cognitive control, such as response inhibition, in non-clinical populations (48). Large-scale evidence has also shown that the right IFG and median cingulate are critically involved in the cognitive re-appraisal of emotional stimuli (49). The reduced activity found in the right IFG and median cingulate in schizophrenia are consistent with positive and negative emotional phenotypes observed in these patients.

Amygdala activity was found to be unaltered in schizophrenia. This negative result contrasts with previous studies, which have reported both hypo- and hyper-activations of the amygdala in schizophrenia (50,51). It is therefore possible that opposite results cancelled out each other, especially since the SDM-PSI approach is based on effect size estimates, regardless of their directionality. Furthermore, population characteristics, such as positive symptoms, negative symptoms, and emotional problems have all been demonstrated to impact amygdala findings in schizophrenia (52). Finally, amygdala alterations in schizophrenia may vary based on specific task contrasts. The amygdala activity would be weakly blunted in response to negative stimuli, and significantly increased in response to ambiguous stimuli, as shown by our research team (51). Here, analyses were performed using all contrasts available for all the emotional tasks that were used. This approach makes the amygdala results in ASD even more compelling as they were robust to methodological differences across studies.

While the emotional paradigms produced robust results, the analyses on functional neuroimaging studies using social cognition tasks (mostly theory of mind) produced no significant results using corrected statistical thresholds. This is somewhat surprising considering that schizophrenia and ASD are associated with notable deficits in social cognition, including deficits in theory of mind, with large effect sizes in most cases (53). Studies using the quantitative theory of mind performance to discriminate between schizophrenia and ASD have generally failed to show significant differences between both disorders (54). Methodological issues may have limited the capacity to detect such differences, though (55). Alternatively, the hypo-hyper intentionality hypothesis has proposed that schizophrenia and autism may be differentiated qualitatively based on the mechanisms involved in theory of mind deficits in both disorders (27). According to this model, the over-attribution of intentions to other people in schizophrenia (hyper-intentionality) would contrast with a relative lack of spontaneous attribution of mental states to others (hypo-intentionality) in ASD. Using an uncorrected threshold, ASD individuals showed reduced activation in the right superior temporal gyrus, while schizophrenia patients displayed reduced activity in the right (posterior) middle temporal gyrus. Although both brains regions are known to be involved in several similar socio-cognitive functions (56,57), these results were not sufficiently robust to help establish commonalities or differences between schizophrenia and ASD.

When examining only studies on mental state attribution, we observed neural alterations in ASD individuals specifically, in the right posterior insula as well as in the right superior temporal gyrus. Noteworthy, the right middle temporal gyrus was also found to be altered in ASD in the global analysis on social cognition studies (see Supplementary Table 8 for the overlap), meaning that this latter result was mostly driven by the studies using mental state attribution tasks. As for the posterior insula, it is well-known to play a significant role in the perception of physiological states. The alteration found in this region may therefore suggest that the deficits in mental state attribution in ASD individuals may arise (in part) from an improper interpretation of their own bodily-related experiences (50). However, these results should be interpreted cautiously since they were based on only 16 studies.

The main strength of the current meta-analysis is that it has included a substantially larger number of studies than the previous meta-analysis from Sugranyes et al. (28) on socio-emotional processing in schizophrenia and ASD. While the previous meta-analysis had included only 33 studies, our meta-analysis has included a total of 184 functional neuroimaging studies, meaning that we had considerably more statistical power to investigate both commonalities and differences between schizophrenia and ASD. Another strength of the meta-analysis is that the main results obtained with the SDM-PSI software were replicated using ALE. This attests to the robustness of results.

Despite these strengths, our meta-analysis suffers from a few limitations. The first limitation has to do with the sociodemographic profiles of individuals with both disorders. Since autism is typically diagnosed in childhood and psychosis onset typically occurs in late adolescence or early adulthood, age differences may have theoretically biased our results. Thus, we excluded studies in ASD individuals younger than 18 years old and schizophrenia patients older than 40 to make sure that the age range of studies was identical for both disorders. Differences in sex ratio were observed between disorders, with a lower ratio of females in ASD, which may have partially accounted for some of our results. However, we performed meta-regression analyses using sex ratio as a moderator variable and found no relationship with any findings. The fact that schizophrenia patients were receiving antipsychotic treatment is another limitation. Again, we performed meta-regression analyses, and found no association between neural alterations and chlorpromazine equivalents. Finally, despite including a large number of studies in the meta-analysis, the sample size of each study was generally small (n<30), meaning that some of the included studies had reduced power to detect differences between clinical and non-clinical populations.

While the search for common traits and neural alterations in schizophrenia and autism is gaining favor, the current meta-analysis showed a lack of spatial conjunction of the neural mechanisms involved in emotion processing in both disorders. During emotion processing, ASD was associated with alterations of bottom-up mechanisms (amygdala), while schizophrenia was associated, to the opposite, with alterations of top-down mechanisms (right IFG), suggesting that it is possible to differentiate both disorders at the neural level. Differences in neuroimaging pattern and developmental trajectories of socio-emotional behavioral components in the two conditions become evident when examined with a higher granularity than ‘’traits’’(21). As the evidence gathered here was not the product of a direct comparison between autism spectrum and schizophrenia, future functional neuroimaging studies will need to perform head-to-head comparisons of patients with schizophrenia and ASD, while using sophisticated emotional paradigms. Such investigations will need to use machine learning approaches to determine the classification accuracy of the task-based functional neuroimaging results. Careful attention will need to be paid to the definition of inclusion criteria, since it has been shown that the adoption of liberal diagnostic criteria impedes the ability to detect brain alterations in autism (14).

## Data Availability

All data referred to in this study is available in the current literature.

## Acknowledgments

SP is holder of the Eli Lilly Canada Chair on schizophrenia research; LM is holder of the Marcel and Rolande Gosselin Research Chair in Cognitive Neurosciences of Autism; MB is holder of a doctoral scholarship from the Canadian Institutes of Health Research (FBD-193359). FP is holder of a Merit scholarship from the University of Montreal.

## Conflicts of interest

None to declare.

## Sources of funding

None

Supplementary information is available at MP’s website.

## Notes

### Competing Interest Statement

The authors have declared no competing interest.

### Funding Statement

This study did not receive any funding

