## Supplementary Material for "Distinct neural alterations in schizophrenia and autism: A meta-analysis of social cognition and emotion processing"

**Figure S1:** Flow chart of study selection for a functional neuroimaging meta-analysis during a social cognition/emotional task in patients with schizophrenia

Records from EMBASE (n= 1096)

Records from Web of Science (n= 1353)

Records from PubMed (n= 1339)

All records (n= 3788)

Records screened (n= 1776)

Records removed from duplicates (n= 2012)

Records rejected based on abstract (n= 1612)

Reports excluded for various reasons (n= 60):

- No non-clinical group (n= 1)

- Missing data or no between-group comparison (n= 17)

- Predefined regions of interest (n= 29)

- Task is not related (n = 3)

- >40 years old (n= 10)

Reports for eligibility (n= 164)

Studies included (n= 104)

**Figure S2:** Flow chart of study selection for a functional neuroimaging meta-analysis during a social cognition/emotional task in patients with autism spectrum disorder

Records from EMBASE (n= 840)

Records from Web of Science (n= 1033)

Records from PubMed (n= 963)

All records (n= 2836)

Records screened (n= 1712)

Records removed from duplicates (n= 1124)

Records rejected based on abstract (n= 1488)

Reports excluded for various reasons (n= 144):

- <18 years old (n= 57)

- Missing data or no between-group comparison (n= 31)

- Predefined regions of interest (n= 41)

- Task is not related (n = 15)

Reports for eligibility (n= 224)

Studies included (n= 80)

**Table S1.** PRISMA checklist

| Section/topic | # | Checklist item | Reported on page # |
| --- | --- | --- | --- |
| TITLE | | |  |
| Title | 1 | Identify the report as a systematic review, meta-analysis, or both. | 1 |
| ABSTRACT | | |  |
| Structured summary | 2 | Provide a structured summary including, as applicable: background; objectives; data sources; study eligibility criteria, participants, and interventions; study appraisal and synthesis methods; results; limitations; conclusions and implications of key findings; systematic review registration number. | 3 |
| INTRODUCTION | | |  |
| Rationale | 3 | Describe the rationale for the review in the context of what is already known. | 3 |
| Objectives | 4 | Provide an explicit statement of questions being addressed with reference to participants, interventions, comparisons, outcomes, and study design (PICOS). | 3 |
| METHODS | | |  |
| Protocol and registration | 5 | Indicate if a review protocol exists, if and where it can be accessed (e.g., Web address), and, if available, provide registration information including registration number. | --- |
| Eligibility criteria | 6 | Specify study characteristics (e.g., PICOS, length of follow-up) and report characteristics (e.g., years considered, language, publication status) used as criteria for eligibility, giving rationale. | 6-7 |
| Information sources | 7 | Describe all information sources (e.g., databases with dates of coverage, contact with study authors to identify additional studies) in the search and date last searched. | 6 |
| Search | 8 | Present full electronic search strategy for at least one database, including any limits used, such that it could be repeated. | 6 |
| Study selection | 9 | State the process for selecting studies (i.e., screening, eligibility, included in systematic review, and, if applicable, included in the meta-analysis). | 6-7 |
| Data collection process | 10 | Describe method of data extraction from reports (e.g., piloted forms, independently, in duplicate) and any processes for obtaining and confirming data from investigators. | 7 |
| Data items | 11 | List and define all variables for which data were sought (e.g., PICOS, funding sources) and any assumptions and simplifications made. | --- |
| Risk of bias in individual studies | 12 | Describe methods used for assessing risk of bias of individual studies (including specification of whether this was done at the study or outcome level), and how this information is to be used in any data synthesis. | 8 |
| Summary measures | 13 | State the principal summary measures (e.g., risk ratio, difference in means). | 8 |
| Synthesis of results | 14 | Describe the methods of handling data and combining results of studies, if done, including measures of consistency (e.g., I^2^) for each meta-analysis. | 8-9 |
| Risk of bias across studies | 15 | Specify any assessment of risk of bias that may affect the cumulative evidence (e.g., publication bias, selective reporting within studies). | 8 |
| Additional analyses | 16 | Describe methods of additional analyses (e.g., sensitivity or subgroup analyses, meta-regression), if done, indicating which were pre-specified. | 8-10 |

| RESULTS | | |  |
| --- | --- | --- | --- |
| Study selection | 17 | Give numbers of studies screened, assessed for eligibility, and included in the review, with reasons for exclusions at each stage, ideally with a flow diagram. | 10 |
| Study characteristics | 18 | For each study, present characteristics for which data were extracted (e.g., study size, PICOS, follow-up period) and provide the citations. | 10 and supplementary material |
| Risk of bias within studies | 19 | Present data on risk of bias of each study and, if available, any outcome level assessment (see item 12). |  |
| Results of individual studies | 20 | For all outcomes considered (benefits or harms), present, for each study: (a) simple summary data for each intervention group (b) effect estimates and confidence intervals, ideally with a forest plot. | --- |
| Synthesis of results | 21 | Present results of each meta-analysis done, including confidence intervals and measures of consistency. | 11-13 |
| Risk of bias across studies | 22 | Present results of any assessment of risk of bias across studies (see Item 15). | 11-12 |
| Additional analysis | 23 | Give results of additional analyses, if done (e.g., sensitivity or subgroup analyses, meta-regression [see Item 16]). | 11-12 |
| DISCUSSION | | |  |
| Summary of evidence | 24 | Summarize the main findings including the strength of evidence for each main outcome; consider their relevance to key groups (e.g., healthcare providers, users, and policy makers). | 13 |
| Limitations | 25 | Discuss limitations at study and outcome level (e.g., risk of bias), and at review-level (e.g., incomplete retrieval of identified research, reporting bias). | 17-18 |
| Conclusions | 26 | Provide a general interpretation of the results in the context of other evidence, and implications for future research. | 14-16 |
| FUNDING | | |  |
| Funding | 27 | Describe sources of funding for the systematic review and other support (e.g., supply of data); role of funders for the systematic review. | 18 |

From: Moher D, Liberati A, Tetzlaff J, Altman DG, The PRISMA Group (2009). Preferred Reporting Items for Systematic Reviews and Meta-Analyses: The PRISMA Statement. PLoS Med 6(7): e1000097. doi:10.1371/journal.pmed1000097

For more information, visit: www.prisma-statement.org.

| **Table S2.** Characteristics of the studies on schizophrenia | | | | | | | | |
| --- | --- | --- | --- | --- | --- | --- | --- | --- |
|  |  |  |  |  |  | **PANSS** | |  |
| **Study** | **n patients** | **n controls** | **Mean age** | **% of females** | **Cpz dose** | **Positive** | **Negative** | **Ratio SCZ** |
| (Andreasen et al., 2008) | 18 | 13 | 32 | 27,78 | 0 | 11,91 | 7,83 | 100 |
| (Bartholomeusz et al., 2018) | 14 | 22 | 20,43 | 42,86 | 1146,58 |  | 15,86 | 100 |
| (Blain et al., 2023) | 62 | 54 | 32,9 | 51,61 | 330 | 17 | 14,6 | 35 |
| (Brüne et al., 2008) | 9 | 13 | 27,89 | 66,67 | 244,44 | 16,67 | 15,67 | 100 |
| (Brüne et al., 2011) | 22 | 26 | 26,8 | 36,36 | 475 | 18,2 | 21,2 | 100 |
| (Brunet et al., 2003) | 7 | 8 | 31,4 | 0 | 709,43 | 17,8 | 19,5 | 100 |
| (Ciaramidaro et al., 2018) | 20 | 25 | 24,7 | 30 |  | 14,2 | 17,4 | 100 |
| (Comte et al., 2018) | 26 | 26 | 32,31 | 34,62 | 274,36 | 8,04 | 13,04 | 100 |
| (Dar et al., 2021) | 31 | 17 | 34,7 | 19,4 | 559,7 | 2,2 | 2,1 | 97 |
| (Das et al., 2012) | 23 | 22 | 34,5 | 0 |  | 10,1 | 18,2 | 100 |
| (De Coster et al., 2019) | 23 | 25 | 35,3 | 0 | 908,49 | 11,56 | 12,13 |  |
| (Derntl et al., 2012) | 15 | 25 | 34,2 | 33,33 | 329,9 | 12,3 | 14,6 | 100 |
| (Dodell-Feder et al., 2014) | 20 | 18 | 38,8 | 40 | 501,6 | 3,1 | 1,7 | 80 |
| (Dollfus et al., 2008) | 23 | 23 | 29,9 | 21,7 | 315,5 | 11,7 | 13,1 | 100 |
| (Dowd & Barch, 2010) | 40 | 32 | 36,8 | 35 | 452,2 | 11,66 | 7,73 |  |
| (Dyck et al., 2014) | 16 | 16 | 35,94 | 37,5 |  | 10,13 | 12,69 | 100 |
| (Ebisch et al., 2013) | 24 | 22 | 27,3 | 33,33 | 422 | 13,1 | 12 | 100 |
| (Fakra et al., 2008) | 14 | 14 | 37,29 | 35,71 |  | 24,71 | 13,14 | 100 |
| (Fakra et al., 2008) | 22 | 22 | 37,29 | 35,71 |  | 24,71 | 13,14 | 100 |
| (Ferri et al., 2014) | 26 | 30 | 27,45 | 36,36 | 480 | 12,54 | 12,04 | 100 |
| (Garcia-Leon et al., 2021) | 12 | 12 | 38,46 | 7,69 |  | 15,88 | 22,46 |  |
| (Gizewski et al., 2013) | 13 | 16 | 37,8 | 0 | 672,3 | 14,6 | 20,5 | 100 |
| (Gur et al., 2002) | 14 | 14 | 28,8 | 28,57 | 472,13 | 11,32 | 7,66 | 100 |
| (Gur et al., 2007) | 20 | 20 | 30,1 | 25 | 652,29 | 11,55 | 7,56 | 100 |
| (Habel, Pauly, et al., 2010) | 14 | 14 | 37,14 | 0 |  | 19,42 | 23,5 | 100 |
| (Habel, Chechko, et al., 2010) | 17 | 17 | 34,4 |  |  | 18 | 19,9 | 100 |
| (J. Hall et al., 2008) | 24 | 24 | 35,1 | 36,84 | 496 | 12,3 | 11,8 | 100 |
| (He et al., 2021) | 17 | 18 | 33,12 | 23,53 | 562,52 | 15,07 | 14,18 | 93,33 |
| (Herold et al., 2018) | 12 | 12 | 36,88 | 50 |  | 14 | 18,02 | 100 |
| (Holt et al., 2012) | 20 | 17 | 34,7 | 0 | 301 | 13,5 | 13,9 | 100 |
| (Horne et al., 2022) | 30 | 40 | 26,8 | 30 | 242,2 | 18,7 | 16,1 | 100 |
| (Kang et al., 2009) hallucinator | 14 | 28 | 29,5 | 50 | 701,8 | 17,9 | 17,3 | 100 |
| (Kang et al., 2009) non-hallucinator | 14 | 28 | 30,4 | 50 | 435,6 | 13,8 | 16,1 | 100 |
| (Kosaka et al., 2002) | 12 | 12 | 26 | 50 | 322 | 11,3 | 16,3 | 100 |
| (Lakis et al., 2011) | 37 | 37 | 32,46 | 49 | 613,92 | 18,84 | 12,59 | 100 |
| (K.-H. Lee et al., 2006) | 14 | 14 | 31,7 | 7 | 354,3 | 12,97 | 9,67 | 100 |
| (S. J. Lee et al., 2010) | 15 | 18 | 26 | 53,33 | 422,1 | 13,1 | 15,4 | 100 |
| (J. Lee et al., 2011) | 14 | 14 | 38,3 | 20 |  |  |  | 100 |
| (S.-K. Lee et al., 2014) | 15 | 16 | 36,7 | 40 | 454,5 | 9,7 | 12,6 | 100 |
| (J. S. Lee et al., 2014) | 15 | 14 | 31,7 | 46,7 | 489,1 | 13,4 | 15,8 | 100 |
| (Leitman et al., 2011) | 23 | 28 | 34,1 | 34,8 | 416,2 | 12,51 | 9,34 |  |
| (Lemmers-Jansen et al., 2019) | 26 |  | 19,88 | 36,36 |  | 13,23 | 17,18 | 100 |
| (Li et al., 2012) | 12 | 12 | 29,8 | 50 | 404,87 | 16,08 | 13,41 | 100 |
| (Linnman et al., 2013) | 15 | 15 | 32 | 0 | 341 | 14 | 13 | 100 |
| (Makowski et al., 2016) | 15 | 15 | 33,1 | 73,33 |  | 14,27 | 13,74 | 100 |
| (Mendrek et al., 2012) | 22 | 25 | 32,86 | 100 | 496,61 | 19,32 | 20,14 | 100 |
| (Michalopoulou et al., 2008) | 11 | 9 | 35 | 18,18 | 523 | 16 | 13,91 | 100 |
| (Mier et al., 2010) | 16 | 16 | 34,25 | 31,25 | 901,59 | 11,58 | 9,79 | 100 |
| (Mier et al., 2014) | 12 | 16 | 32,45 | 34 | 472,56 | 11,56 | 9,34 | 100 |
| (Mukherjee et al., 2014) | 24 | 20 | 35,13 | 40 | 494 | 12,3 | 15,8 | 100 |
| (Oh et al., 2015) | 16 | 16 | 29,8 | 50 | 311,1 | 17,6 | 17,7 | 100 |
| (Okruszek et al., 2018) | 23 | 26 | 35,7 | 48 | 332 | 11,4 | 18,4 | 100 |
| (Paradiso et al., 2003) | 18 | 17 | 30 | 11,11 | 0 | 11,84 | 7,97 | 100 |
| (K.-M. Park et al., 2009) | 15 | 16 |  |  | 410 | 13,1 | 17,1 | 100 |
| (Pedersen et al., 2012) | 15 | 14 | 29 | 40 | 629,6 | 10,9 | 14,9 | 100 |
| (Pinkham et al., 2011) | 35 | 37 | 36,46 | 51,4 | 378,19 | 16,88 | 14,18 | 88,57 |
| (Pinkham et al., 2018) | 31 | 32 | 35,65 | 42 | 452,26 |  | 12,23 | 38,71 |
| (Quarto et al., 2018) | 40 | 56 | 33,2 | 40 | 536 |  |  | 100 |
| (Rapp et al., 2013) | 15 | 15 | 28,1 | 100 | 516 | 17,4 | 16 | 100 |
| (Regenbogen et al., 2015) | 20 | 24 | 37,3 |  |  | 14,21 | 23,11 | 100 |
| (Reske et al., 2007) | 10 | 10 | 37,4 | 40 | 207,68 | 8,9 | 16,1 | 100 |
| (Russell et al., 2000) | 5 | 7 | 36 | 0 |  |  |  | 100 |
| (Schnell et al., 2016) | 21 | 24 |  | 57,1 |  |  |  | 100 |
| (Shin et al., 2015) | 16 | 16 | 32 | 0 |  | 12,1 | 16,5 | 100 |
| (Soldevila-Matías et al., 2023) Chronic | 23 | 31 | 34,7 | 52,1 |  | 16,83 | 17,26 | 100 |
| (Soldevila-Matías et al., 2023) FEP | 31 | 31 | 28,97 | 25,8 |  | 16,42 | 16,19 |  |
| (Stegmayer et al., 2018) | 22 | 25 | 37,5 | 36 | 397,5 | 17,5 | 18,8 | 100 |
| (Straube et al., 2013) | 16 | 16 | 38 | 37,5 | 601,25 | 16 | 16 | 100 |
| (Szabó et al., 2017) | 19 | 18 | 37,6 | 33,33 | 716 | 17,6 | 20,3 | 100 |
| (Takahashi et al., 2004) | 15 | 15 | 29 | 33,33 |  |  |  | 100 |
| (Taylor et al., 2011) | 21 | 21 | 40,7 | 33,33 |  |  | 13,13 | 77 |
| (Ursu et al., 2011) | 23 | 24 | 29,4 | 25 |  | 12,04 | 8,38 | 91 |
| (Varga et al., 2013) | 21 | 24 | 37,95 | 57,1 |  | 13,81 | 17 | 100 |
| (Walter et al., 2009) | 12 | 12 | 29,5 | 50 |  | 17,75 | 19,41 | 100 |
| (L. (Lea) M. Williams et al., 2007) non paranoid | 14 | 13 | 27,8 | 35,7 | 339,3 | 13,5 | 18,6 | 100 |
| (L. (Lea) M. Williams et al., 2007) paranoid | 13 | 13 | 26,9 | 38,46 | 375,1 | 24,8 | 22,4 | 100 |
| (Anticevic et al., 2011) | 28 | 24 | 36,39 | 22 | 584,63 | 11,68 | 7,96 | 100 |
| (Berger et al., 2018) | 31 | 19 | 32,6 | 29 | 493,77 | 13,91 | 10,5 | 73 |
| (Bliksted et al., 2014) | 17 | 17 | 23,94 | 29,4 |  | 14,99 | 10,4 | 100 |
| (Briend et al., 2019) | 20 | 28 | 39,69 | 35 | 296,91 | 12,5 | 11,8 | 100 |
| (Diaz et al., 2011) | 11 | 17 | 32,57 | 9 |  | 15 | 18 | 100 |
| (Guimond et al., 2018) | 20 | 20 | 26,9 | 40 | 511,7 | 13,38 | 13,97 | 100 |
| (Kim et al., 2015) | 15 | 15 | 28,4 | 53 |  | 15,4 | 18,1 | 100 |
| (Kohler et al., 2008) | 11 | 10 | 35,4 | 54 |  |  |  | 100 |
| (Larabi et al., 2018) | 30 | 15 | 35 | 27 |  | 14,47 | 14,27 | 100 |
| (J. Lee et al., 2016) | 15 | 14 | 36,7 | 40 | 454,5 |  |  | 100 |
| (H. Lee et al., 2014) | 15 | 16 | 30,7 | 33 | 499 | 16,2 | 15,4 | 100 |
| (Lindner et al., 2014) | 36 | 40 | 30,8 | 38,8 |  | 12,69 | 10,48 | 100 |
| (Mukerji et al., 2018) | 19 | 24 | 38,1 | 47 | 350,88 | 16,37 | 13,05 | 78,9 |
| (J.-I. Park et al., 2019) | 17 | 17 | 31,1 | 47 |  | 16,5 | 19,5 | 100 |
| (Rahm et al., 2015) | 28 | 28 | 29,9 | 25 |  |  | 13,9 | 100 |
| (Razafimandimby et al., 2016) | 21 | 25 | 33,9 | 23,8 | 340,3 | 13,6 | 12 | 100 |
| (Satterthwaite et al., 2010) | 16 | 21 |  | 40 | 290 |  |  | 75 |
| (Sergerie et al., 2010) | 20 | 20 | 31,8 | 45 | 377 | 13,44 | 12,5 | 100 |
| (Singh et al., 2015) | 14 | 14 | 31,5 | 21,4 | 389,3 | 13,41 | 11,36 | 100 |
| (Smith et al., 2015) | 30 | 24 | 33,6 | 40 | 362,8 | 13,78 | 12,2 | 100 |
| (Spilka et al., 2019) | 23 | 26 | 40 | 30 |  | 14,6 | 14 | 65 |
| (Swart et al., 2013) | 18 | 18 | 29,44 | 16,6 |  |  |  | 100 |
| (Taylor et al., 2005) | 18 | 10 | 32,47 | 38,8 | 400,12 |  | 9,75 | 100 |
| (Tikàsz et al., 2016) | 20 | 21 | 30 | 0 | 846,9 | 9,1 | 12,9 |  |
| (H.-H. Tseng et al., 2016) | 18 | 21 | 27,72 | 27,7 | 186,66 | 13,47 | 13,17 |  |
| (Vercammen et al., 2012) | 20 | 23 | 34,4 | 25 |  | 15,9 | 16,1 | 80 |
| (Villalta-Gil et al., 2013) | 22 | 31 | 23,34 | 40,9 | 390,2 | 16,83 | 18,02 |  |
| (Vistoli et al., 2017) | 27 | 21 | 29,7 | 14,8 | 547,7 | 13,8 | 16 | 67,67 |
| (L. M. Williams et al., 2004) | 27 | 22 | 27,3 | 37 | 356,5 | 19,47 | 20,3 | 100 |
| (Whalley et al., 2009) | 15 | 14 | 38,4 | 26,6 |  |  |  | 100 |
| (Adamczyk et al., 2017) | 20 | 20 | 39,95 | 50 | 495 | 10,6 | 14,95 | 80 |
| (Schiffer et al., 2017) | 16 | 18 | 38,4 | 0 | 599 | 7 | 17,4 | 100 |
| Note. CPZ = Chlorpromazine; PANSS = Positive and Negative Syndrome Scale | | | | | | | | |

| **Table S3.** Characteristics of the studies on autism spectrum disorder | | | | |
| --- | --- | --- | --- | --- |
| **Study** | **n patients** | **n controls** | **age** | **% of females** |
| (Alaerts et al., 2014) | 15 | 15 | 21,7 | 0 |
| (Antezana et al., 2022) | 15 | 16 | 26,14 | 13,33 |
| (Aoki et al., 2014) | 17 | 17 | 29,6 | 0 |
| (Baron‐Cohen et al., 1999) | 6 | 12 | 26,3 | 33,33 |
| (Bird et al., 2010) | 18 | 18 | 34,6 | 0 |
| (Bölte et al., 2015) | 32 | 25 | 19,3 | 6,25 |
| (Caria et al., 2011) | 8 | 12 | 24,3 | 25 |
| (Castelli, 2002) | 10 | 10 | 33 |  |
| (Charpentier et al., 2020) | 14 | 16 | 27,9 | 7,14 |
| (Chen et al., 2021) | 26 | 25 | 19,5 | 0 |
| (Ciaramidaro et al., 2018) | 33 | 25 | 18,78 | 6 |
| (Corradi-Dell’Acqua et al., 2014) | 10 | 10 | 21,5 | 0 |
| (Critchley et al., 2000) | 9 | 9 | 37 | 0 |
| (Daly et al., 2012) | 14 | 14 | 31 | 0 |
| (Deeley et al., 2007) | 9 | 9 | 34 | 0 |
| (Dufour et al., 2013) | 27 | 27 | 31 | 18,5 |
| (Fan et al., 2014) | 24 | 21 | 18,4 | 0 |
| (Fittipaldi, Armony, García, et al., 2023) | 30 | 27 | 28,8 | 50 |
| (Fittipaldi, Armony, Migeot, et al., 2023) | 30 | 28 | 28,8 | 50 |
| (Gebauer, Skewes, Westphael, et al., 2014) | 19 | 20 | 26,16 | 10,5 |
| (Gebauer, Skewes, Hørlyck, et al., 2014) | 19 | 20 | 26,16 | 10,5 |
| (Georgescu et al., 2013) | 13 | 13 | 31,23 | 30,76 |
| (Graves et al., 2022) | 19 | 22 | 20,8 | 21 |
| (Grèzes et al., 2009) | 12 | 12 | 27 | 16,67 |
| (Gu et al., 2015) | 17 | 17 | 26,2 | 0 |
| (Hadjikhani et al., 2009) | 9 | 7 | 30 | 25 |
| (Hadjikhani et al., 2014) | 36 | 31 | 24 | 8 |
| (G. B. C. Hall et al., 2003) | 8 | 8 | 26,5 | 0 |
| (Happé et al., 1996) | 5 | 6 | 24 | 0 |
| (Hsu et al., 2018) | 26 | 30 | 35,08 | 42 |
| (Ishitobi et al., 2011) | 9 | 24 | 23 | 11,1 |
| (Ilzarbe et al., 2020) | 19 | 19 | 23 | 0 |
| (Kana et al., 2009) | 12 | 12 | 24,6 | 16,6 |
| (Kana et al., 2014) | 15 | 15 | 21 |  |
| (Kennedy & Courchesne, 2008) | 12 | 14 | 26,5 | 0 |
| (Kirkovski et al., 2016) | 27 | 23 | 30,56 | 51,8 |
| (Kliemann et al., 2018) | 30 | 50 | 28,27 | 23,3 |
| (Kuzmanovic et al., 2014) | 13 | 13 | 29,08 | 38,4 |
| (Lassalle et al., 2017) | 27 | 21 | 24 | 0 |
| (Lassalle et al., 2019) | 19 | 20 | 25,27 | 10,5 |
| (Lee Masson et al., 2020) | 21 | 21 | 25 | 0 |
| (Libero et al., 2014) | 27 | 23 | 21,04 | 14,8 |
| (Loveland et al., 2008) | 5 | 4 | 18 | 20 |
| (Marsh & Hamilton, 2011) | 18 | 19 | 33 |  |
| (Mason et al., 2008) | 18 | 18 | 26,5 | 5,55 |
| (Moseley et al., 2015) | 18 | 18 | 30,4 |  |
| (Murdaugh et al., 2012) | 13 | 14 | 21,4 | 0 |
| (Nijhof et al., 2018) | 24 | 21 | 32,8 | 45,8 |
| (Pantelis et al., 2015) | 17 | 21 | 28,9 | 29,4 |
| (Pelphrey et al., 2007) | 8 | 8 | 25 | 25 |
| (Perlman et al., 2011) | 12 | 7 | 25,5 | 8,3 |
| (Pierce, 2004) | 8 | 10 | 27,1 | 0 |
| (Pitskel et al., 2011) | 15 | 14 | 23,4 | 0 |
| (Procyshyn et al., 2022) | 16 | 21 | 29,9 | 100 |
| (Richey et al., 2015) | 15 | 15 | 26,14 | 13,3 |
| (Richey et al., 2022) | 21 | 20 | 21,62 | 0 |
| (Rosenblau, Kliemann, Lemme, et al., 2016) | 27 | 22 | 33 | 35,7 |
| (Rosenblau, Kliemann, Dziobek, et al., 2016) | 27 | 22 | 33 | 33,33 |
| (Salmi et al., 2013) | 13 | 13 | 29 | 0 |
| (Sato et al., 2012) | 31 | 13 | 27,2 | 8,33 |
| (Sato et al., 2019) | 12 | 31 | 27,5 | 29,03 |
| (Schneider, Pauly, et al., 2013) | 28 | 28 | 31 | 46,43 |
| (Schneider, Regenbogen, et al., 2013) females | 13 | 13 | 29,85 | 100 |
| (Schneider, Regenbogen, et al., 2013) males | 15 | 15 | 32,73 | 0 |
| (Schulte-Rüther et al., 2011) | 14 | 18 | 27 | 0 |
| (Schulte-Rüther et al., 2011) | 14 | 18 | 27 | 0 |
| (Shafritz et al., 2015) | 15 | 18 | 18 | 20 |
| (Silani et al., 2008) | 15 | 15 | 37 | 13,33 |
| (Sommer et al., 2018) | 15 | 15 | 28,2 | 33,33 |
| (Stanfield et al., 2017) | 28 | 33 | 40 | 21,43 |
| (Stroth et al., 2019) | 9 | 9 | 18,7 | 100 |
| (Tam et al., 2017) | 22 | 24 | 34,1 | 0 |
| (Tanabe et al., 2012) | 21 | 21 | 25,1 | 23,81 |
| (A. Tseng et al., 2016) | 51 | 84 | 27,5 | 12 |
| (Velasquez et al., 2017) | 19 | 22 | 26 | 31,58 |
| (Vinckier et al., 2021) | 19 | 19 | 26,8 | 31,58 |
| (Watanabe et al., 2012) | 15 | 17 | 28,2 | 0 |
| (Zürcher et al., 2013) | 16 | 22 | 23,5 | 13,64 |
| (Chanel et al., 2016) | 15 | 14 | 28,6 | 13,33 |
| (Pelphrey et al., 2005) | 10 | 9 | 23,2 | 10 |
| (Sato et al., 2017) | 16 | 17 | 26,1 | 6,25 |
| (Wicker et al., 2008) | 12 | 14 | 27 | 8,33 |

| **Table S4.** Classification of Autism spectrum diagnosis and phenotype. | | | | | | | | | |
| --- | --- | --- | --- | --- | --- | --- | --- | --- | --- |
|  |  |  |  |  |  |  | **Clinical judgement** | |  |
| **Study** | **Dx** | **Dx criteria** | **Phenotype** | **% autism** | **ADOS** | **ADI-R** | **1** | **2** | **Other details** |
| (Alaerts et al., 2014) | Autism | DSM-IV | High | 100 | X |  |  |  | Previous Dx. Parental Social Responsiveness Scale. Recruited from a “center”. |
| (Antezana et al., 2022) | ASD | NS | Low | 33,33 | X |  |  |  | Previous Dx. Recruited from Autism Subject Registry of an Institute. |
| (Aoki et al., 2014) | HFA | DSM-IV-TR | Low | 58,82 | X | X (did not meet threshold) |  |  | Maternal reported Social Responsiveness Scale. Recruited from outpatient clinic |
| (Baron‐Cohen et al., 1999) | Autism & Asperger | DSM-IV & ICD-10 | Low | NS |  |  |  |  | Recruitment not specified. |
| (Bird et al., 2010) | Autism & Asperger | DSM-IV | Low | 16,67 |  |  |  |  | Recruitment not specified. |
| (Bölte et al., 2015) | Autism, Asperger & PDD-NOS | ICD-10 | Low | 31,25 | X | X |  |  | Recruitment not specified. |
| (Caria et al., 2011) | Asperger | DSM-IV & ICD-10 | Low | 0 | X |  |  |  | Recruited through Internet advertisement. |
| (Castelli, 2002) | Autism & Asperger | DSM-IV | Low | NS |  |  |  |  | Recruitment not specified. |
| (Charpentier et al., 2020) | ASD | DSM-IV-TR | Low | NS | X | X |  | X | Recruited from a “center”. |
| (Chen et al., 2021) | ASD | DSM-V | Low | NS |  |  |  |  | Recruited from a community autism program. |
| (Ciaramidaro et al., 2018) | Autism, Asperger & atypical autism | ICD-10 | Low | 30,3 | X | X |  | X | Recruited from community. |
| (Corradi-Dell’Acqua et al., 2014) | HFA & Asperger | NS | Low | 40 | X | X |  |  | Recruited from database of a specialized clinic for PDD |
| (Critchley et al., 2000) | Autism & Asperger | ICD-10 | Low | 22,2 |  | X |  |  | Recruitment not specified. |
| (Daly et al., 2012) | Autism | ICD-10 | High | 100 |  | X |  |  | Childhood Dx.  Recruited through clinical research program. |
| (Deeley et al., 2007) | Asperger | DSM-IV & ICD-10 | Low | 0 | X | X |  |  | Recruitment not specified. |
| (Dufour et al., 2013) | ASD | NS | Low | NS | X |  |  |  | Recruitment not specified. |
| (Fan et al., 2014) | ASD | DSM-IV | Low | NS |  | X |  |  | Recruitment not specified. |
| (Fittipaldi, Armony, García, et al., 2023) | Autism & ASD | DSM-V | Low | NS | X |  | X |  | Recruitment not specified. |
| (Fittipaldi, Armony, Migeot, et al., 2023) | Autism & ASD | DSM-V | Low | NS | X |  | X |  | Recruitment not specified. |
| (Gebauer, Skewes, Westphael, et al., 2014) | Autism & Asperger | NS | Low | NS |  |  |  |  | Recruited through the national autism and Asperger’s association. |
| (Gebauer, Skewes, Hørlyck, et al., 2014) | Autism & Asperger | NS | Low | NS |  |  |  |  | Recruited through the national autism and Asperger’s association. |
| (Georgescu et al., 2013) | HFA & Asperger | ICD-10 | Low | NS |  |  |  | X | Recruited from outpatient clinic. |
| (Graves et al., 2022) | ASD | NS | Low | NS | X |  | X |  | Previous Dx. Recruited by advertisement and word-of-mouth contacts. |
| (Grèzes et al., 2009) | Autism & Asperger | DSM-IV | Low | 16,67 |  |  |  |  | Recruitment not specified. |
| (Gu et al., 2015) | Autism & Asperger | DSM-IV-TR | Low | 70,5 | X | X |  |  | Recruited from a “center”. |
| (Hadjikhani et al., 2009) | Autism, Asperger & PDD-NOS | DSM-IV | Low | 27,78 | X | X |  |  | Recruited from “centers”. |
| (Hadjikhani et al., 2014) | Autism, Asperger & PDD-NOS | DSM-IV | Low | 27,78 | X | X |  |  | Recruited from “centers”. |
| (G. B. C. Hall et al., 2003) | Autism & Asperger | DSM-IV | Low | 75 |  |  |  |  | Recruitment not specified. |
| (Happé et al., 1996) | Asperger | NS | Low | 0 |  |  |  |  | Previous Dx. Recruitment not specified |
| (Hsu et al., 2018) | ASD | DSM-IV | Low | NS | X |  |  |  | Previous Dx. Recruited from a database of research volunteers with and without ASD |
| (Ishitobi et al., 2011) | HFA & Asperger | DSM-IV | Low | 33,33 |  |  | X |  | Diagnosed based on the Diagnostic Interview for Social and Communication Disorders (DISCO). Recruited from hospital. |
| (Ilzarbe et al., 2020) | Autism, Asperger, atypical autism & PDD-NOS | ICD-10 | Low | 44,44 | X | X |  |  | 14 with previous Dx and 5 with a research Dx. Recruitment through specialists, support organizations, social media, and a database. |
| (Kana et al., 2009) | HFA | NS | High | 100 | X | X | X |  | Recruitment not specified. |
| (Kana et al., 2014) | Asperger & ASD | NS | Low | 46,67 | X | X |  |  | Previous Dx. Recruited from clinics. |
| (Kennedy & Courchesne, 2008) | HFA, Asperger & PDD-NOS | NS | Low | 20 | X | X | X |  | Recruitment not specified. |
| (Kirkovski et al., 2016) | Asperger & autism | DSM-IV | Low | 14,8 |  |  |  |  | >30% female. Previous Dx verified. |
| (Kliemann et al., 2018) | ASD | NS | Low | NS | X |  |  |  | Previous Dx. |
| (Kuzmanovic et al., 2014) | Asperger & autism | ICD-10 | Low | NS |  |  |  | X | >30% female. Recruited from the outpatient clinic for autism. |
| (Lassalle et al., 2017) | Asperger, ASD & PDD-NOS | DSM-IV-TR | Low | 22,22 | X | X |  |  | Recruitment in three cities but not specified how. |
| (Lassalle et al., 2019) | Asperger, ASD & PDD-NOS | DSM-IV | Low | 31,58 | X | X |  |  | Recruitment in two cities but not specified how. |
| (Lee Masson et al., 2020) | HFA | DSM-IV or DSM-5 | Low | NS |  |  |  |  | Previous Dx. Adult Asperger Assessment Inventory was also administered. |
| (Libero et al., 2014) | HFA | NS | High | 100 | X | X |  |  | Previous Dx. Recruitment not specified. |
| (Loveland et al., 2008) | Autism | DSM-IV | High | 100 | X | X |  |  | Recruitment not specified. |
| (Marsh & Hamilton, 2011) | Asperger & ASD | NS | Low | 50 | X |  |  |  | Recruitment not specified. |
| (Mason et al., 2008) | HFA | NS | High | 100 | X | X | X |  | Recruitment not specified. |
| (Moseley et al., 2015) | Asperger & PDD-NOS | DSM-IV | Low | 0 |  |  |  |  | Previous Dx. Recruitment through the database for autism research. |
| (Murdaugh et al., 2012) | Autism & Asperger | NS | Low | 53,85 | X | X | X |  | Recruitment through the local community and the university database. |
| (Nijhof et al., 2018) | HFA | NS | Low | 70,83 | X |  | X |  | >30% female. Previous Dx. 7 scored below the ADOS cut-off. Recruitment through advertising. |
| (Pantelis et al., 2015) | ASD | DSM-IV-TR | High | 100 | X | X | X |  | Recruitment not specified. |
| (Pelphrey et al., 2007) | HFA | NS | High | 100 | X | X | X |  | History of clinical diagnosis. Recruited from a center for research on autism. |
| (Perlman et al., 2011) | Autism | DSM-IV | High | 100 | X | X | X |  | Previous Dx. Recruitment not specified. |
| (Pierce, 2004) | Autism | DSM-IV | High | 100 | X | X |  |  | Recruited from the hospital. |
| (Pitskel et al., 2011) | Autism | DSM-IV | High | 100 | X | X | X |  | Previous dx. No Asperger or PDD-NOS dx. |
| (Procyshyn et al., 2022) | Autistic & Asperger | DSM-IV & ICD-10 | Low | NS |  |  |  |  | Previous Dx. Recruited from the area. |
| (Richey et al., 2015) | ASD & Asperger | NS | Low | 33,33 | X |  |  |  | History of clinical Dx. Recruited from a registry. |
| (Richey et al., 2022) | ASD | NS | Low | NS | X |  |  |  | Recruitment through flyers in the local  community (e.g., churches, schools, restaurants), existing research registry databases,  university-affiliated assessment clinics, and local ASD support groups. |
| (Rosenblau, Kliemann, Lemme, et al., 2016) | Asperger & autism | DSM-IV | Low | NS | X | X |  |  | >30% female. Recruited from outpatient clinic. |
| (Rosenblau, Kliemann, Dziobek, et al., 2016) | Asperger & HFA | DSM-IV | Low | NS | X | X |  |  | Also administered to Asperger the Asperger Syndrome and  High-Functioning Autism Diagnostic Interview. Recruited from outpatient clinic. |
| (Salmi et al., 2013) | Asperger | ICD-10 | Low | 0 |  |  | ? |  | Diagnosis process included a detailed developmental history. Recruitment not specified. |
| (Sato et al., 2012) | Asperger & PDD-NOS | DSM-IV-TR | Low | 0 |  |  |  | X | Administered the Childhood Autism Rating Scale. Recruitment not specified. |
| (Sato et al., 2019) | Asperger & PDD-NOS | DSM-IV-TR | Low | 0 |  |  |  | X | Administered the Childhood Autism Rating Scale. Recruitment not specified. |
| (Schneider, Pauly, et al., 2013) | HFA | DSM-IV | Low | NS | X |  |  |  | >30% female. Pre or complete previous Dx. Recruited from in/outpatient facilities, local self-help groups and therapy centers. |
| (Schneider, Regenbogen, et al., 2013) females | Autistic | DSM-IV | Low | 100 | X |  |  | X | >30% female. Pre or complete previous Dx. Recruited from in/outpatient facilities, local self-help groups and therapy centers. 7 did not fulfilled ADOS criteria, therefore additional information from relatives and therapists were collected. |
| (Schneider, Regenbogen, et al., 2013) males | Autistic | DSM-IV | High | 100 | X |  |  | X | Pre or complete previous Dx. Recruited from in/outpatient facilities, local self-help groups and therapy centers. 7 did not fulfilled ADOS criteria, therefore additional information from relatives and therapists were collected. |
| (Schulte-Rüther et al., 2011) | HFA & Asperger | ICD-10 & DSM-IV | Low | 50 | X |  |  | ? | Recruitment not specified. |
| (Shafritz et al., 2015) | Autistic disorder & Asperger | DSM-IV | Low | 73,33 | X | X |  |  | Recruited from a center for autism. |
| (Silani et al., 2008) | HFA & Asperger | DSM-IV | Low | NS | X |  |  |  | Previous Dx. Recruitment not specified. |
| (Sommer et al., 2018) | Autism & Asperger | ICD-10 | Low | 26,67 |  |  |  | ? | >30% female. Recruited from outpatient services. |
| (Stanfield et al., 2017) | Autism & Asperger | DSM-IV | Low | NS | X |  |  |  | Recruited from clinical services. |
| (Stroth et al., 2019) | Asperger & atypical autism | DSM-IV & ICD-10 | Low | 0 | X | X |  |  | >30% female. Recruited from outpatient clinic. |
| (Tam et al., 2017) | ASD | NS | High | 100 | X |  |  |  | Previous Dx. Recruitment not specified. |
| (Tanabe et al., 2012) | HFA, autism & Asperger | DSM-IV-TR | Low | 76,19 |  |  |  | X | DISCO (Diagnostic Interview for Social and Communication Disorders). Recruited from hospitals. |
| (A. Tseng et al., 2016) | Autism, Asperger & PDD-NOS | DSM-IV-TR | Low | 35,29 | X | X | X |  | Recruited from the area. |
| (Velasquez et al., 2017) | ASD | DSM-IV | Low | 100 | X | X | ? |  | >30% female |
| (Vinckier et al., 2021) | Autism | DSM-IV | Low | 100 |  |  |  | ? | >30% female, recruited from the clinic for autism. |
| (Watanabe et al., 2012) | HFA & PDD-NOS | DSM-IV |  | 93,33 |  | X |  | X | “For all participants who did not meet the threshold  in the ADI-R social domain, the group was confirmed by the  Childhood Autism Rating Scale (CARS)”. Recruited from outpatient service. |
| (Zürcher et al., 2013) | HFA | DSM-IV & DSM-5 | High | 100 | X | X |  | ? | Recruited from “centers” |
| (Chanel et al., 2016) | ASD | DSM-IV |  |  | X |  |  |  | Previous Dx. Recruitment not specified. |
| (Pelphrey et al., 2005) | Autism |  |  |  | X | X |  |  | Previous Dx. Recruited through clinical institutes. |
| (Sato et al., 2017) | Asperger & PDD-NOS | DSM-IV & DSM-5 | Low | 0 |  |  |  | X | Interviews with parents and participants. Recruitment not specified. |
| (Wicker et al., 2008) | Autism & Asperger | DSM-IV | Low | 66,67 |  |  |  |  | “Parents of all the subjects were asked to answer the screening questionnaire for autistic spectrum disorders”  Recruited from associations of parents |
| Note. ? = unclear if there were two clinicians in the consensus for the diagnosis or only one; ADOS = Autism Diagnostic Observation Schedule; ADI-R = Autism Diagnostic Interview, Revised; NS = Not specified; ASD = autism spectrum disorder; HFA = high-functioning autism; PDD-NOS = pervasive developmental disorder not otherwise specified; DSM = Diagnostic and Statistical Manual; ICD = International Classification of Diseases | | | | | | | | | |

| **Table S5.** Paradigms included in the meta-analyses | | | |
| --- | --- | --- | --- |
|  |  | **Classification** | |
| **Study** | **Name of the task** | **Emotion** | **Social cognition** |
| **Schizophrenia** | | | |
| (Andreasen et al., 2008) | Theory of mind story |  | **X** |
| (Bartholomeusz et al., 2018) | Picture-story attribution-of-intentions theory of mind |  | **X** |
| (Blain et al., 2023) | Eye gaze processing |  | **X** |
| (Brüne et al., 2008) | Cartoon-based theory of mind |  | **X** |
| (Brüne et al., 2011) | Cartoon-based theory of mind |  | **X** |
| (Brunet et al., 2003) | Comic strips theory of mind |  | **X** |
| (Ciaramidaro et al., 2018) | Facial affect recognition | **X** |  |
| (Comte et al., 2018) | Variable attention and congruency task | **X** |  |
| (Dar et al., 2021) | Emotional word paradigm | **X** |  |
| (Das et al., 2012) | Theory of mind (geometric shapes) |  | **X** |
| (De Coster et al., 2019)a | False belief |  | **X** |
| (De Coster et al., 2019)b | Person description |  | **X** |
| (De Coster et al., 2019)c | Emotional theory of mind | **X** |  |
| (Derntl et al., 2012)a | Emotion recognition | **X** |  |
| (Derntl et al., 2012)b | Emotional perspective taking | **X** |  |
| (Derntl et al., 2012)c | Affective responsiveness | **X** |  |
| (Dodell-Feder et al., 2014) | False belief |  | **X** |
| (Dollfus et al., 2008) | Listening task of a story with characters and social interactions |  | **X** |
| (Dowd & Barch, 2010) | valence and arousal ratings of emotional pictures (words, pictures and faces) | **X** |  |
| (Dyck et al., 2014) | Audiovisual mood induction | **X** |  |
| (Ebisch et al., 2013) | Social perception |  | **X** |
| (Fakra et al., 2008)a | Intuitive emotional condition (matching emotional faces) | **X** |  |
| (Fakra et al., 2008)b | Cognitively demanding condition (labeling emotional faces) | **X** |  |
| (Ferri et al., 2014) | Emotional goal-related actions | **X** |  |
| (Garcia-Leon et al., 2021) | Emotional pictures task | **X** |  |
| (Gizewski et al., 2013) | Reading the mind in the eyes |  | **X** |
| (Gur et al., 2002) | Valence judgment and facial emotion processing | **X** |  |
| (Gur et al., 2007) | Facial emotion identification task | **X** |  |
| (Habel, Pauly, et al., 2010) | Emotion was induced by odorants during an n-back working memory task | **X** |  |
| (Habel, Chechko, et al., 2010) | Emotional discrimination | **X** |  |
| (J. Hall et al., 2008) | Passive view of facial expressions | **X** |  |
| (He et al., 2021) | Social vs. non-social videos to identify neural perception of social and non-social information in both auditory-speech and visual-gesture modalities |  | **X** |
| (Herold et al., 2018) | Irony comprehension |  | **X** |
| (Holt et al., 2012) | Fear conditioning and extinction | **X** |  |
| (Horne et al., 2022) | Reward learning task | **X** |  |
| (Kang et al., 2009) | Processing of laugh and crying sounds | **X** |  |
| (Kosaka et al., 2002) | Emotional intensity judgment task. | **X** |  |
| (Lakis et al., 2011) | Emotional recognition memory | **X** |  |
| (K.-H. Lee et al., 2006) | Empathic and forgivability judgements |  | **X** |
| (S. J. Lee et al., 2010) | Cognitive empathy |  | **X** |
| (S. J. Lee et al., 2010) | Emotional empathy | **X** |  |
| (S. J. Lee et al., 2010) | Inhibitory empathy |  | **X** |
| (J. Lee et al., 2011) | False belief |  | **X** |
| (S.-K. Lee et al., 2014) | Emotional salience attribution | **X** |  |
| (J. S. Lee et al., 2014) | Pre-trained facial expression task | **X** |  |
| (Leitman et al., 2011) | Four-choice (happiness, fear, anger, neutral) vocal affect identification task | **X** |  |
| (Lemmers-Jansen et al., 2019) | Trust game |  | **X** |
| (Li et al., 2012) | Facial emotion perception | **X** |  |
| (Li et al., 2012) | Fear conditioning | **X** |  |
| (Linnman et al., 2013) | Social approval task | **X** |  |
| (Makowski et al., 2016) | Social approval task | **X** |  |
| (Mendrek et al., 2012) | Emotion-Processing Task | **X** |  |
| (Michalopoulou et al., 2008) | Facial fear processing | **X** |  |
| (Mier et al., 2010)a | Affective theory of mind |  | **X** |
| (Mier et al., 2010)b | Processing of faces with neutral expressions | **X** |  |
| (Mier et al., 2010)c | Emotion recognition | **X** |  |
| (Mier et al., 2014) | Adaptive emotion recognition task | **X** |  |
| (Mukherjee et al., 2014) | Approachability judgement |  | **X** |
| (Oh et al., 2015) | Theme-identification task | **X** |  |
| (Okruszek et al., 2018) | Recognition of communicative interactions |  | **X** |
| (Paradiso et al., 2003) | Emotion attribution protocol | **X** |  |
| (K.-M. Park et al., 2009) | Attributional task | **X** |  |
| (Pedersen et al., 2012) | “Moving Shapes” paradigm |  | **X** |
| (Pinkham et al., 2011) | Emotion recognition task | **X** |  |
| (Pinkham et al., 2018) | Social cognitive introspective accuracy (i.e., emotion recognition) | **X** |  |
| (Quarto et al., 2018) | Emotion processing | **X** |  |
| (Rapp et al., 2013) | Irony comprehension |  | **X** |
| (Regenbogen et al., 2015) | Empathy assessment | **X** |  |
| (Reske et al., 2007) | Mood induction using facial expressions | **X** |  |
| (Russell et al., 2000) | Mental state attribution |  | **X** |
| (Schnell et al., 2016) | Irony comprehension |  | **X** |
| (Shin et al., 2015) | Reading the mind in the eyes |  | **X** |
| (Soldevila-Matías et al., 2023) | Emotional and nonemotional words | **X** |  |
| (Stegmayer et al., 2018) | Gesture planning and execution |  | **X** |
| (Straube et al., 2013) | Processing of metaphoric gestures |  | **X** |
| (Szabó et al., 2017) | Recognition of mixed emotions | **X** |  |
| (Takahashi et al., 2004) | Affective pictures | **X** |  |
| (Taylor et al., 2011) | Social appraisal | **X** |  |
| (Ursu et al., 2011) | Emotional experience task | **X** |  |
| (Varga et al., 2013) | Irony comprehension |  | **X** |
| (Walter et al., 2009) | Theory of mind |  | **X** |
| (L. (Lea) M. Williams et al., 2007) | Passive view of facial expressions | **X** |  |
| (Anticevic et al., 2011) | A modified delayed-response visual working memory task faced with affectively negative, neutral, or task-related interference | **X** |  |
| (Berger et al., 2018) | Humor processing |  | **X** |
| (Bliksted et al., 2014) | Theory of mind |  | **X** |
| (Briend et al., 2019) | Language fMRI task (infer mental states) |  | **X** |
| (Diaz et al., 2011) | Verbal working memory task with emotional distraction | **X** |  |
| (Guimond et al., 2018) | Emotional n-back (faces) | **X** |  |
| (Kim et al., 2015) | Implicit memory task with emotional words | **X** |  |
| (Kohler et al., 2008) | Eye gaze discrimination |  | **X** |
| (Larabi et al., 2018) | Emotion regulation | **X** |  |
| (J. Lee et al., 2016) | Emotion attribution task (judge) | **X** |  |
| (J. Lee et al., 2016) | Emotion attribution task (view) | **X** |  |
| (H. Lee et al., 2014) | Emotional salience attribution | **X** |  |
| (J. Lee et al., 2016) | Handshake: acceptance vs refusal processing | **X** |  |
| (Lindner et al., 2014) | Facial disgust | **X** |  |
| (Mukerji et al., 2018) | Simulation fMRI task | **X** |  |
| (J.-I. Park et al., 2019) | A delayed response working memory task that included neutral and fearful distractors | **X** |  |
| (Rahm et al., 2015) | Affective processing task | **X** |  |
| (Razafimandimby et al., 2016) | Emotional sentence attribution | **X** |  |
| (Satterthwaite et al., 2010) | Facial recognition memory | **X** |  |
| (Sergerie et al., 2010) | Modulation of emotion on memory recognition | **X** |  |
| (Singh et al., 2015) | Empathy task | **X** |  |
| (Smith et al., 2015) | Cognitive empathy task | **X** |  |
| (Spilka et al., 2019) | Facial emotion perception task | **X** |  |
| (Swart et al., 2013) | Associative emotional learning | **X** |  |
| (Taylor et al., 2005) | Neurobehavioral probe with emotional and nonemotional stimuli | **X** |  |
| (Tikàsz et al., 2016) | Emotion processing task | **X** |  |
| (H.-H. Tseng et al., 2016) | Facial and prosodic emotional recognition | **X** |  |
| (Vercammen et al., 2012) | Emotional go-nogo (words) | **X** |  |
| (Villalta-Gil et al., 2013) | Explicit and implicit processing of fearful and happy facial expressions | **X** |  |
| (Vistoli et al., 2017) | Perspective taking task | **X** |  |
| (L. M. Williams et al., 2004) | Pictures depicting facial expressions of fear or neutral emotion | **X** |  |
| (Whalley et al., 2009) | Emotional memory | **X** |  |
| (Adamczyk et al., 2017) | Humor processing |  | **X** |
| (Schiffer et al., 2017) | Affective theory of mind |  | **X** |
| **Autism spectrum disorder** | | | |
| (Alaerts et al., 2014) | Emotion recognition task | **X** |  |
| (Antezana et al., 2022) | Emotion regulation | **X** |  |
| (Aoki et al., 2014) | First-order false belief task (emotional state attribution) | **X** |  |
| (Aoki et al., 2014) | First-order false belief task (belief attribution) |  | **X** |
| (Baron‐Cohen et al., 1999) | Reading the mind in the eyes |  | **X** |
| (Bird et al., 2010) | Empathy for pain task | **X** |  |
| (Bölte et al., 2015) | Explicit facial affect recognition | **X** |  |
| (Bölte et al., 2015) | Implicit facial affect recognition | **X** |  |
| (Caria et al., 2011) | Emotional processing of music | **X** |  |
| (Castelli, 2002) | Attribution of mental states to animated shapes |  | **X** |
| (Charpentier et al., 2020) | Oddball with emotional stimuli (prosody) | **X** |  |
| (Chen et al., 2021) | Explicit and implicit (backwardly masked) perception of threatening faces | **X** |  |
| (Ciaramidaro et al., 2018) | Explicit facial affect recognition | **X** |  |
| (Ciaramidaro et al., 2018) | Implicit facial affect recognition | **X** |  |
| (Corradi-Dell’Acqua et al., 2014) | Passive viewing of emotional faces | **X** |  |
| (Critchley et al., 2000) | Processing of emotional facial expressions. | **X** |  |
| (Daly et al., 2012) | Incidental processing of disgust, fearful, happy, and sad facial expressions | **X** |  |
| (Deeley et al., 2007) | Facial Emotion Processing | **X** |  |
| (Dufour et al., 2013) | False belief task |  | **X** |
| (Fan et al., 2014) | Empathy-eliciting stimuli depicting physical bodily injuries | **X** |  |
| (Fittipaldi, Armony, García, et al., 2023) | Intent-based moral judgement |  | **X** |
| (Fittipaldi, Armony, Migeot, et al., 2023) | Envy task | **X** |  |
| (Gebauer, Skewes, Westphael, et al., 2014) | Emotion of affective prosody | **X** |  |
| (Gebauer, Skewes, Hørlyck, et al., 2014) | Processing of musical emotions | **X** |  |
| (Georgescu et al., 2013) | Eye gaze processing |  | **X** |
| (Graves et al., 2022) | Social and figurative language processing |  | **X** |
| (Grèzes et al., 2009) | Viewing actions with and without an emotional meaning | **X** |  |
| (Gu et al., 2015) | Empathy-for-pain task | **X** |  |
| (Hadjikhani et al., 2009) | Perception of bodily expressed emotions | **X** |  |
| (Hadjikhani et al., 2014) | Emotional contagion for pain task | **X** |  |
| (G. B. C. Hall et al., 2003) | Emotion attribution | **X** |  |
| (Happé et al., 1996) | Theory of mind |  | **X** |
| (Hsu et al., 2018) | Mimic faces task | **X** |  |
| (Ishitobi et al., 2011) | Processing whole faces and parts of faces displaying positive or negative expressions | **X** |  |
| (Ilzarbe et al., 2020) | Frith-Happé animated-triangle theory of mind task |  | **X** |
| (Kana et al., 2009) | Theory of mind (geometric figures) |  | **X** |
| (Kana et al., 2014) | Theory of mind (comic strips) |  | **X** |
| (Kennedy & Courchesne, 2008) | Emotional stroop task (words) | **X** |  |
| (Kirkovski et al., 2016) | Animation task |  | **X** |
| (Kliemann et al., 2018) | Dynamic emotional facial expressions | **X** |  |
| (Kuzmanovic et al., 2014) | Animacy rating |  | **X** |
| (Lassalle et al., 2017) | Viewing pictures of neutral faces and faces expressing anger, happiness, and fear at low and high intensity, with a fixation cross between the eyes | **X** |  |
| (Lassalle et al., 2019) | Perception of others’ pain | **X** |  |
| (Lee Masson et al., 2020) | Positive versus negative affective touch processing | **X** |  |
| (Libero et al., 2014) | Attribution of Emotions to Body Postures | **X** |  |
| (Loveland et al., 2008) | Judgments of auditory-visual affective congruence | **X** |  |
| (Marsh & Hamilton, 2011) | Rational and irrational hand actions processing task |  | **X** |
| (Mason et al., 2008) | Narrative comprehension | **X** |  |
| (Mason et al., 2008) | Inference task |  | **X** |
| (Moseley et al., 2015) | Abstract emotion word processing | **X** |  |
| (Murdaugh et al., 2012) | Theory of mind |  | **X** |
| (Nijhof et al., 2018) | Explicit and spontaneous mentalizing |  | **X** |
| (Pantelis et al., 2015) | Quantify social awkwardness |  | **X** |
| (Pelphrey et al., 2007) | Dynamic perception of facial affect and identity | **X** |  |
| (Perlman et al., 2011) | Fearful faces processing | **X** |  |
| (Pierce, 2004) | Familiarity and the perception of face identity | **X** |  |
| (Pitskel et al., 2011) | Eye gaze processing |  | **X** |
| (Procyshyn et al., 2022) | Processing emotional faces vs shapes | **X** |  |
| (Richey et al., 2015) | Emotion regulation | **X** |  |
| (Richey et al., 2022) | Facial emotion recognition | **X** |  |
| (Rosenblau, Kliemann, Lemme, et al., 2016) | Implicit naturalistic mentalizing task |  | **X** |
| (Rosenblau, Kliemann, Dziobek, et al., 2016) | Emotional prosody processing | **X** |  |
| (Salmi et al., 2013) | Viewing a feature film portraying social interactions | **X** |  |
| (Sato et al., 2012) | Processing of dynamic and static facial expressions of fear and happiness | **X** |  |
| (Sato et al., 2019) | Processing of facial expression stimuli | **X** |  |
| (Schneider, Pauly, et al., 2013) | Moral decision making |  | **X** |
| (Schneider, Regenbogen, et al., 2013) | Processing of self-related short stories. | **X** |  |
| (Schulte-Rüther et al., 2011) | Identify the emotional state observed or felt | **X** |  |
| (Shafritz et al., 2015) | Faces go/no-go | **X** |  |
| (Silani et al., 2008) | Rate stimuli according to the degree of un/pleasantness | **X** |  |
| (Sommer et al., 2018) | False belief task |  | **X** |
| (Stanfield et al., 2017) | Approachability judgment task |  | **X** |
| (Stroth et al., 2019) | Empathy for pain | **X** |  |
| (Tam et al., 2017) | n-back with emotional faces | **X** |  |
| (Tanabe et al., 2012) | Eye gaze exchange |  | **X** |
| (A. Tseng et al., 2016) | Ratings of arousal and valence for a broad range of emotional faces | **X** |  |
| (Velasquez et al., 2017) | Faces go/no-go | **X** |  |
| (Vinckier et al., 2021) | Rating likeability of ambiguous faces | **X** |  |
| (Watanabe et al., 2012) | Friend or foe judgments of realistic movies | **X** |  |
| (Zürcher et al., 2013) | Eye gaze processing |  | **X** |
| (Chanel et al., 2016) | Emotional facial/body expression | **X** |  |
| (Pelphrey et al., 2005) | Eye gaze processing |  | **X** |
| (Sato et al., 2017) | Eye gaze processing |  | **X** |
| (Wicker et al., 2008) | Explicit emotion processing | **X** |  |

| **Table S6.** Frequency of paradigms according to disorders. | | | |
| --- | --- | --- | --- |
| **Categories** | **Type of paradigms** | **Schizophrenia** | **Autism spectrum disorder** |
| *Social cognition* | | | |
| Inference/Mental state attribution | - Theory of mind - False-belief task / belief attribution - Reading the mind in the eyes - Cognitive empathy - Trust game - Intent-based moral judgement - Mentalizing task | 21 (17.6%) | 16 (18.6%) |
| Eye gaze processing | - Averted/directed eye gaze processing | 2 (1.7%) | 6 (7.0%) |
| Social | - Person description - Listening task with social interactions - Social perception - Empathic and forgivability judgement - Viewing gesture and/or hand actions - Animacy rating - Quantify social awkwardness - Approachability judgement | 9 (7.6%) | 4 (4.7%) |
| Double meaning | - Social and figurative language processing - Humor/irony processing | 6 (5.0%) | 1 (1.2%) |
| *Emotion* | | | |
| Explicit | - Rate pleasantness - Identify emotion - Matching of facial expressions | 47 (39.5%) | 35 (40.7%) |
| Implicit | - Passive view of emotional content - Gender discrimination task | 34 (28.6%) | 24 (28.0%) |
| Note. The distribution of paradigms did not differ between disorders according to whether it was an inference task (theory of mind task, false belief task, reading the mind in the eyes, etc.) (X^2^(1, 65)=0.103, p=0.749), another social cognition task (eye gaze processing, approachability judgement, humor processing, etc.) (X^2^(1, 192)=0.005, p=0.945) or an explicit/implicit emotional task (X^2^(1, 155)=0.001, p=0.973) | | | |

| **Table S7**. Seed-based d mapping uncorrected results of lower activations in schizophrenia compared to non-clinical controls. | | | | | |
| --- | --- | --- | --- | --- | --- |
| Emotional tasks | | | | | |
| **MNI coordinates** | **SDM-Z** | **p-value** | **Voxels** | **Peak** | **Cluster breakdown** |
| 50,32,16 | -7.222 | ~0 | 1241 | R inferior frontal gyrus, triangular part | R inferior frontal gyrus triangular part, R insula, R IFG orbital part, R middle frontal gyrus, R superior longitudinal fascicles III, R superior temporal gyrus |
| 2,-38,36 | -5.351 | <0.001 | 336 | R median cingulate / paracingulate gyri | R median cingulate / paracingulate gyri, L median cingulate / paracingulate gyri |
| Social cognition tasks | | | | | |
| **MNI coordinates** | **SDM-Z** | **p-value** | **Voxels** | **Peak** | **Cluster breakdown** |
| 54,-54,10 | -4.222 | <0.001 | 112 | R middle temporal gyrus |  |
| Note. MNI = Montreal Neurological Institute; R = Right. | | | | | |

| **Table S8**. Seed-based d mapping uncorrected results of lower activations in autism spectrum disorder compared to non-clinical controls. | | | | | |
| --- | --- | --- | --- | --- | --- |
| Emotional tasks | | | | | |
| **MNI coordinates** | **SDM-Z** | **p-value** | **Voxels** | **Peak** | **Cluster breakdown** |
| -22,-2,-12 | 7.600 | ~0 | 903 | L amygdala | L amygdala, L striatum, L putamen, Anterior commissure, L hippocampus, L parahippocampal gyrus, L pons, L median network cingulum |
| -54,12,20 | -5.312 | <0.001 | 235 | L inferior frontal gyrus, opercular part | L inferior frontal gyrus opercular part, triangular part, L precentral gyrus |
| -40,-56,-18 | -4.327 | <0.001 | 173 | L fusiform gyrus | L fusiform gyrus, L cerebellum lobule VI, L inferior longitudinal fasciculus |
| 20,-4,-30 | -4.243 | <0.001 | 63 | R parahippocampal gyrus |  |
| Social cognition tasks | | | | | |
| **MNI coordinates** | **SDM-Z** | **p-value** | **Voxels** | **Peak** | **Cluster breakdown** |
| 52,-8,-14 | -4.221 | <0.001 | 92 | R superior temporal gyrus | R superior temporal gyrus, R middle temporal gyrus |
| Note. MNI = Montreal Neurological Institute; L = Left; R = Right. | | | | | |

**Figure S3**. Results from the linear regression analyses.
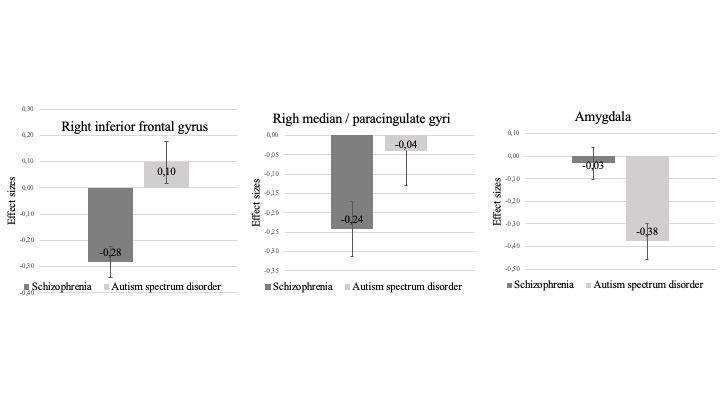

Note. Right inferior frontal gyrus [schizophrenia: g= -0.283 (95 % CI: -0.351 to -0.215)] [autism: g=0.097 (95 % CI: 0.011 to 0.184)]; median cingulate [schizophrenia: g= -0.242 (95 % CI: -310 to -173)] [autism: g= -0.040 (95 % CI: -0.126 to -0.046)]; amygdala [autism: g= -0.376 (95 % CI: -0.463 to -0.289)] [schizophrenia: g= -0.032 (95 % CI: −0.100 to 0.036)]

**Heterogeneity and publication bias for uncorrected results using SDM**

*Emotional tasks in schizophrenia*

There was low heterogeneity (*I*^2^=0.20-10.44%) and no evidence of publication bias (all ps> 0.5). The results were robust as no study affected the meta-analytical estimates by more than 5 %.

*Social cognition tasks in schizophrenia*

Heterogeneity was low (*I*^2^=17.58%) and there was no evidence of publication bias (p=0.824). The result was robust as no study affected the meta-analytical estimates by more than 5 %.

*Emotional tasks in autism*

There was low heterogeneity (*I*^2^=0.20-10.44%) and no evidence of publication bias (all ps> 0.5). The results were robust as no study affected the meta-analytical estimates by more than 5 %.

*Social cognition tasks in autism*

There was low heterogeneity (*I*^2^=4.15%) and no evidence of publication bias (p=0.424). The result was robust as no study affected the meta-analytical estimates by more than 5 %.

*Mental state attribution tasks in autism*

There was low heterogeneity (*I*^2^=0.01%) and no evidence of publication bias (p=0.239). The result was robust as no study affected the meta-analytical estimates by more than 5 %.

**Supplementary Methods: ALE method**

ALE method

The activation likelihood estimation (ALE) approach was used for the coordinated-based meta-analysis (GingerALE version 2.3, https://www. brainmap.org/ale/). This approach allows to test for spatial convergence between studies and requires that peak foci be reported in stereotactic coordinates (Eickhoff et al., 2012). Coordinates of experiments that were reported originally in Talairach stereotaxic space were converted into MNI (Montreal Neurological Institute) space before using them in the analyses. First, a modeled activation map (MA) was created by modeling coordinate foci (x,y,z) with a spherical Gaussian probability distribution, weighted by the number of subjects in each experiment. This is performed to account for spatial uncertainty due to template and between-subject variance (Eickhoff et al., 2009) and to ensure that multiple coordinates from a single experiment do not jointly influence the modeled activation value of a single voxel. Voxel-wise ALE scores were then computed as the union of MA maps, which provide a quantitative assessment of convergence between brain activation across experiments. Then, these maps were cut off by a cluster-forming threshold. In fact, the size of the supra-threshold clusters was compared against a null distribution of cluster sizes derived from artificially created datasets in which foci were shuffled across experiments, but the other properties of original experiments (e.g., number of foci, uncertainty) were kept. Finally, this resulted in calculating the above chance of observing a cluster of the given size (Eickhoff et al., 2012). Consistent with previous meta-analyses, we use the following statistical threshold: a voxel-level cluster forming threshold of p<0.001 and a cluster-level family-wise correction (pFWE < 0.01) (Eickhoff et al., 2016).

Since there was not enough experiments (minimum 17 experiments) to conduct meta-analyses on mental attribution in ASD, these analyses were aborted. On the other hand, there were just enough experiments to conduct a meta-analysis of hyper and hypoactivated focis combined for SCZ studies.

Conjunction analysis in ALE

If significant results were noted in both individual meta-analyses (for example hypoactivation in emotional studies in schizophrenia and autism spectrum disorder), a conjunction analysis was planned. The aim of this analysis was to highlight voxels showing significant activity in both SCZ and ASD during the specific experiment (social cognition and/or emotional paradigms). This analysis is an intersection of ALE maps for each disorder by taking the voxel-wise union of the probability values in order to produce an ALE map (therefore a pooled map). Then, if this pooled map shows significant results, the conjunction analysis can be done. This analysis includes only areas found to be previously significant in ALE maps.

**Supplementary Results: ALE method**

*Emotional paradigms in SCZ*

Included in the analysis for hyperactivated foci are 264 foci,38 experiments and 760 participants with a diagnosis of SCZ. This analysis yielded no significant hyperactivated clusters in emotional paradigms. The analysis for hypoactivated foci included 519 foci, 64 emtoional experiments and 1255 participants with a diagnosis of SCZ which resulted in one significant cluster of hypoactivation in patients compared to HC, namely a brain region encompassing the right anterior insula, inferior frontal gyrus and middle frontal gyrus (Supplementary Table S8).

*Social cognition paradigms in SCZ*

The analysis for hyperactivated foci included 109 foci, 17 experiments and 345 participants with a diagnosis of SCZ which yielded no results at a FWE corrected threshold. The analysis for hypoactivated foci included 172 foci, 30 experiments and 519 participants with a diagnosis of SCZ. This analysis yielded no clusters.

*Mental attribution paradigms in SCZ*

This analysis comprised 169 focis from 21 experiments including 345 participants with a diagnosis of SCZ. No clusters were found for this analysis.

*Emotional paradigms in ASD*

The analysis for hyperactivated foci grouped together 63 foci, 15 experiments and 267 participants with a diagnosis of ASD. No clusters were found with this analysis. The analysis for hypoactivated foci included 315 foci, 38 emotional experiments and 644 clinical participants. This analysis resulted in one significant cluster showing hypoactivation in patients with ASD compared to HC in a brain region encompassing the left amygdala, putamen and para-hippocampal gyrus (Supplementary Table S9).

*Social cognition paradigms in ASD*

Included in the analysis of hyperactivated foci for social cognition paradigms are 52 foci, only 12 experiments (which is not enough to have robust results) and 213 participants with a diagnosis of ASD. This analysis yielded no significant clusters. The analysis for hypoactivated foci included 152 foci, 20 social cognition experiments and 305 clinical participants which resulted in no clusters at a FWE corrected threshold.

*Conjunction analyses*

As can be seen in Supplementary Table 8 and 9, only analyses of hypoactivated foci in emotional paradigms yielded results in both SCZ and ASD. Thus, the only appropriate ALE maps for the conjunction analyses were the ones for the hypoactivated foci in emotional paradigms. We thus performed the pooled analysis to yield an ALE map for both SCZ and ASD. The conjunction analysis revealed no convergence between ASD and SCZ using an uncorrected threshold of p<0.001 or even p<0.05.

| **Table S9.** FWE corrected results with ALE in schizophrenia | | | | | |
| --- | --- | --- | --- | --- | --- |
| Emotional contrasts | | | | | |
| **MNI coordinates** | **ALE value** | **Z-value** | **Cluster size (mm^3^)** | **L/R** | **Cluster breakdown** |
| Hyperactivation | | | | | |
| No clusters found | |  |  |  |  |
| Hypoactivation | | | | | |
| 34,36,-10 | 0.024 | 4.53 | 6184 | R | Insula, inferior frontal gyrus, middle frontal gyrus |
| Social cognition contrasts | | | | | |
| **MNI coordinates** | **ALE value** | **Z-value** | **Cluster size (mm^3^)** | **L/R** | **Cluster breakdown** |
| Hyperactivation | | | | | |
| No clusters found | |  |  |  |  |
| Hypoactivation | | | | | |
| No clusters found | |  |  |  |  |
| Note. FWE = family-wise error; MNI = Montreal Neurological Institute; L = Left; R = Right | | | | | |

| **Table S10.** FWE corrected results with ALE in autism spectrum disorder | | | | | |
| --- | --- | --- | --- | --- | --- |
| Emotional contrasts | | | | | |
| **MNI coordinates** | **ALE value** | **Z-value** | **Cluster size (mm^3^)** | **L/R** | **Cluster breakdown** |
| Hyperactivation | | | | | |
| No clusters found | |  |  |  |  |
| Hypoactivation | | | | | |
| -20,-6,-16 | 0.0298 | 5.94 | 5752 | L | Lentiform nucleus, parahippocampal gyrus, amygdala, putamen, lateral globus pallidus |
| Social cognition contrasts | | | | | |
| **MNI coordinates** | **ALE value** | **Z-value** | **Cluster size (mm^3^)** | **L/R** | **Cluster breakdown** |
| Hyperactivation | | | | | |
| No clusters found | |  |  |  |  |
| Hypoactivation | | | | | |
| No clusters found | |  |  |  |  |
| Note. FWE = family-wise error; MNI = Montreal Neurological Institute; L = Left; R = Right | | | | | |
